# Treatment group outcome variance difference after dropout as an indicator of missing-not-at-random bias in randomized clinical trials

**DOI:** 10.1101/2022.04.15.22273918

**Authors:** Audinga-Dea Hazewinkel, Kate Tilling, Kaitlin H. Wade, Tom Palmer

## Abstract

Randomized controlled trials (RCTs) are considered the gold standard for assessing the causal effect of an exposure on an outcome, but are vulnerable to bias from missing data. When outcomes are missing not at random (MNAR), estimates from complete case analysis (CCA) will be biased. There is no statistical test for distinguishing between outcomes missing at random (MAR) and MNAR, and current strategies rely on comparing dropout proportions and covariate distributions, and using auxiliary information to assess the likelihood of dropout being associated with the outcome. We propose using the observed variance difference across treatment groups as a tool for assessing the risk of dropout being MNAR. In an RCT, at randomization, the distributions of all covariates should be equal in the populations randomized to the intervention and control arms. Under the assumption of homogeneous treatment effects, the variance of the outcome will also be equal in the two populations over the course of followup. We show that under MAR dropout, the observed outcome variances, conditional on the variables included in the model, are equal across groups, while MNAR dropout may result in unequal variances. Consequently, unequal observed conditional group variances are an indicator of MNAR dropout and possible bias of the estimated treatment effect. Heterogeneity of treatment effect affects the intervention group variance, and is another potential cause of observing different outcome variances. We show that, for longitudinal data, we can isolate the effect of MNAR outcome-dependent dropout by considering the variance difference at baseline in the same set of patients that are observed at final follow-up. We illustrate our method in simulation and in applications using individual-level patient data and summary data.

## 1 Introduction

Randomized controlled trials (RCTs) are considered the gold standard for assessing the causal effect of an exposure on an outcome, but are vulnerable to bias due to missingness in the outcome - or ‘dropout’. The impact of dropout depends on the missingness mechanism and the analysis model ^1–3^. Three missingness mechanisms can be distinguished: missing completely at random (MCAR), missing at random (MAR) and missing not at random (MNAR) ^1^. With MCAR, missingness is unrelated to any measured or unmeasured characteristics and the observed sample is a representative subset of the full data. MAR means the missingness can be explained by observed data and, with MNAR, missingness is a function of the unobserved data.

Two common methods of dealing with dropout are complete case analysis (CCA) and multiple imputation (MI). A complete case analysis (CCA) is the analysis model intended to be applied to the trial data at its outset, restricted only to individuals with observed outcomes. With MI, missing outcome values are repeatedly imputed conditional on the observed data, generating multiple complete datasets to which the analysis model is applied ^3–6^, with the resulting estimates subsequently pooled using Rubin’s rules ^4^. Broadly, a CCA will be unbiased under MCAR and MAR dropout, while MI will be unbiased under MCAR and MAR dropout if the imputation model is correctly specified. Generally, both will biased when outcomes are MNAR ^2,3,7^. In this article, we consider the case of an RCT with an incomplete continuous outcome, where the treatment effect is estimated using ordinary least squares (OLS) linear regression. In such an RCT, a CCA will only be biased if the dropout is related the outcome, conditional on the model covariates. ^7–9^

In the presence of dropout, observed data generally cannot be used to establish if outcomes are MNAR or MAR given the model covariates, and consequently whether the complete case treatment effect estimate is likely to be biased. Current guidance for assessing risk of bias due to dropout relies on checking if dropout is differential across treatment groups, assessing the plausibility that dropout may be related to outcome (e.g., dropout due to lack of efficacy) ^10,11^, and comparing the baseline covariate distribution across treatment groups in patients who are still observed at the end of follow-up. ^12^. The European Medical Agency (EMA) and the National Research Council (NRC), recommend using MAR-appropriate methods for the primary analysis, followed by sensitivity analyses that weaken this assumption ^13,14^. These guidelines, however, are in practice implemented in a fraction of all trials, with on average only 6% (of N=330 trials) describing the assumed missing data mechanism ^15,16^, 9% (N=237) justifying their choice of main analysis ^16^, 19% (N=849) reporting some kind of sensitivity analysis ^15–19^, which rarely involves relaxing the primary analysis assumptions ^17^, and only 9% (N=200) discussing the risk of bias resulting from missing data ^19^. This discrepancy between recommended and implemented practice persists despite extensive literature on the subject and may be due to the relative complexity of such analyses.

In this paper, we propose using the observed variances of the outcome in the two arms of the trial to assess the risk of CCA estimator bias due to MNAR dropout. We show, using directed acyclic graphs (DAGs) and standard statistical theory, how MNAR may give rise to unequal outcome variances between the fully-observed subjects in the two arms of the trial. We illustrate this method using individual-level data and summary-level data. Individual-level patient data were obtained from an RCT investigating the benefit of an acupuncture treatment policy for patients with chronic headaches (ISRCTN96537534) ^20^. The summary data application used published statistics from a cluster-randomized clinical trial, which investigated psychological outcomes following a nurse-led preventive psychological intervention for critically ill patients (POPPI, registration ISRCTN53448131).

## 2 Notation

Let *U* be some unmeasured continuous variable that acts on *Y*, with mean *µ*_*U*_ = *γ* and error term *ε*_*U*_, with a mean *µ*_*εU*_ = 0 and variance 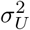:

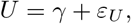

and let *C* be some measured continuous variable, defined analogously:

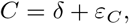

We define the outcome variable, *Y*, as a linear function of treatment, *X* = *j*, where 1 denotes the treatment group and 0 the control group, *U* and *C*, so that

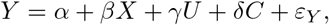

with some intercept, *α, β* the treatment effect, *γ* and *δ* the effects of *U* and *C* on *Y*, respectively, and *ε*_*Y*_ the error term with a mean *µ*_*εY*_ = 0 and variance 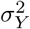.

Let *y*_*ij*_ denote the outcome for a patient *i* (*i* = 1, …, *n*_*j*_) with treatment *j* = 0, 1. Let *µ*_*j*_ denote the population mean for arm *j*, with *µ*_*j*_ = 𝔼 [*Y X* = *j*], estimated by taking the average:

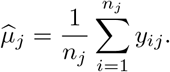

Let *β* denote the true treatment effect, given by the difference in treatment group means, with *β* = *µ*_1_ − *µ*_0_, estimated by

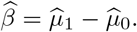

We use ‘full data’ to refer to the data on all members of the population and ‘observed data’ for the sub-sample that is observed. We define a response indicator *R*, with *r*_*ij*_ = 1 when *y*_*ij*_ is observed, and *r*_*ij*_ = 0 when *y*_*ij*_ is missing. Let 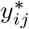 denote the outcome for a patient 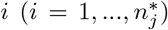 in the observed data, in a given group, *j*, with population mea 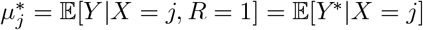, estimated by

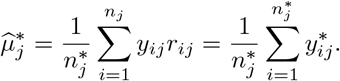

We define the treatment effect in the observed data, 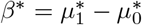, estimated by

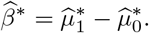

The bias, *B*, of the CCA treatment effect estimate is given by the difference of the population treatment effect in the full data (*β*) and in the observed data (*β*^*\**^):

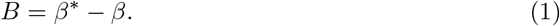

By definition, the population variance of *Y*, for a given treatment arm, *j*, in the full data, is given by

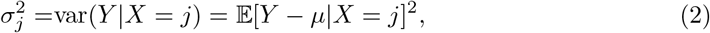

and, in the observed data, by

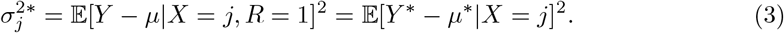

The variance of a given arm, *j*, in the full data (2), is then estimated by

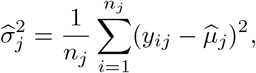

and, in the observed data, (3) by

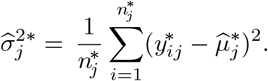

We obtain the unbiased sample variance, 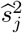 by adjusting 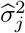^2^ and 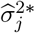 with a factor 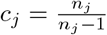 and 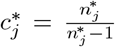 for the full data and observed data, respectively, so that 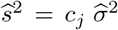 and 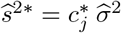

## 3 MAR and MNAR dropout and observed sample variances

In an RCT patients, are randomized to treatment, which makes it plausible to assume that the treatment arms have equal outcome variances in the full data, prior to dropout, or, in the case of repeated measurements, at baseline. Figure 1 shows a flowchart of a two-armed RCT in five phases: (I) the relevant target population is identified; (II) a representative study sample is obtained; (III) patients are randomized to an intervention or control group; (IV) treatment is initiated; (V) patients are followed up, with dropout occurring over the course of follow-up. As patients are randomized to treatment, the population group variances are the same in (III), and will, in the absence of treatment effect heterogeneity, remain the same after treatment initiation in (IV), and over the course of follow-up in (V), given that no dropout occurs. Then, it follows that if dropout is present and the variances are different, this must be due to dropout. Here, we use directed acyclic graphs (DAGs) to describe different MAR and MNAR dropout mechanisms, and show, using standard statistical theory ^21^, that the treatment arm variances, conditional on the model covariates, are the same under an MAR dropout mechanism, but may be different under an MNAR dropout mechanism.

**Figure 1:**
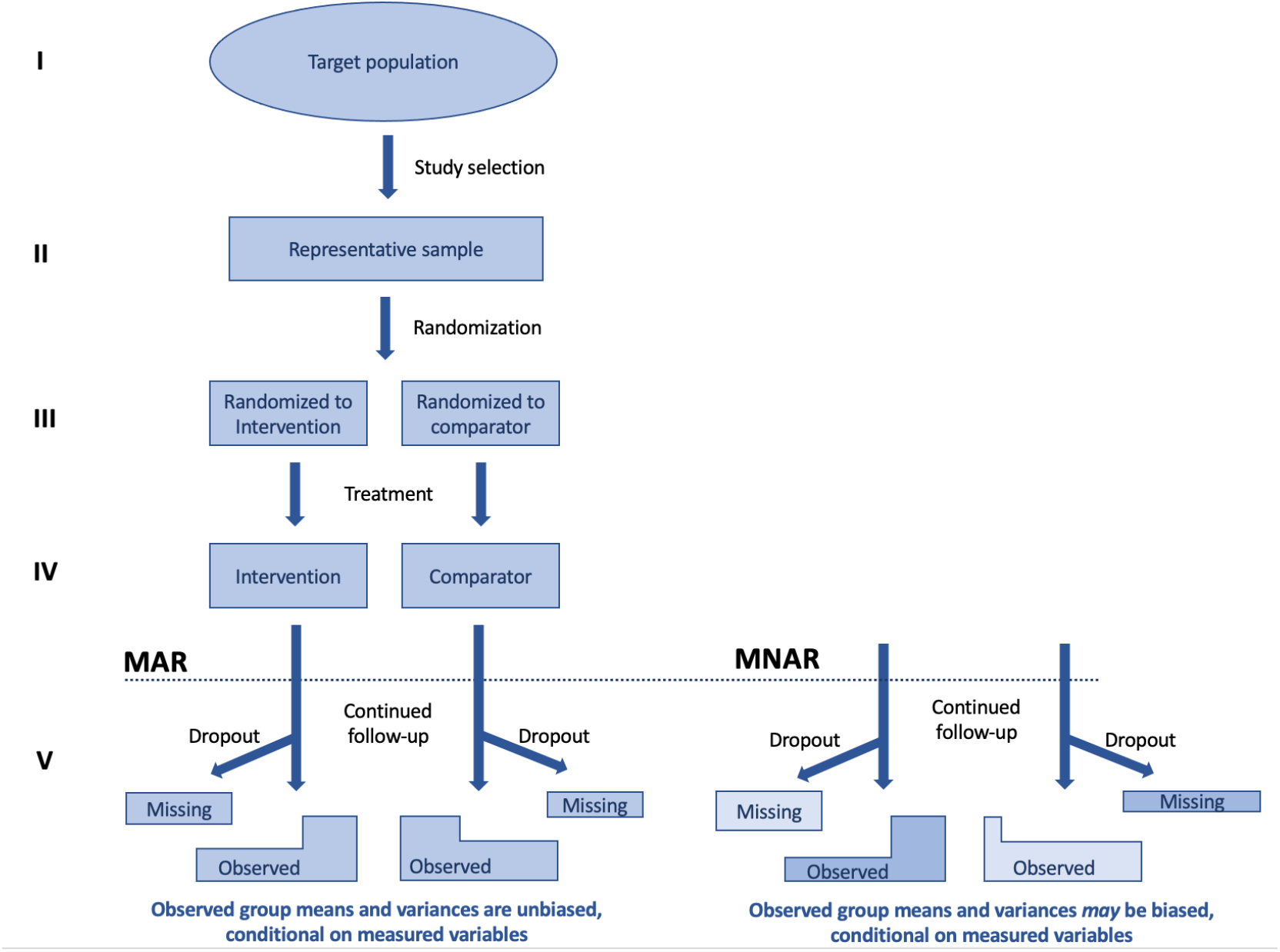
Flowchart of the phases of an RCT with an intervention and control arm: (I) target population; (II) study sample; (III) randomization to intervention/comparator group; (IV) treatment initiation; (V) continued follow-up, subject to patient dropout, which may be MAR (left) or MNAR (right)

### 3.1 Proposition 1

Figure 2.A depicts an MAR dropout mechanism, where *Y* is some function *f* (*X, U*), and *R* some function *g*(*X*). By definition, we can write the joint density of *Y, U*, and *X* as

**Figure 2:**
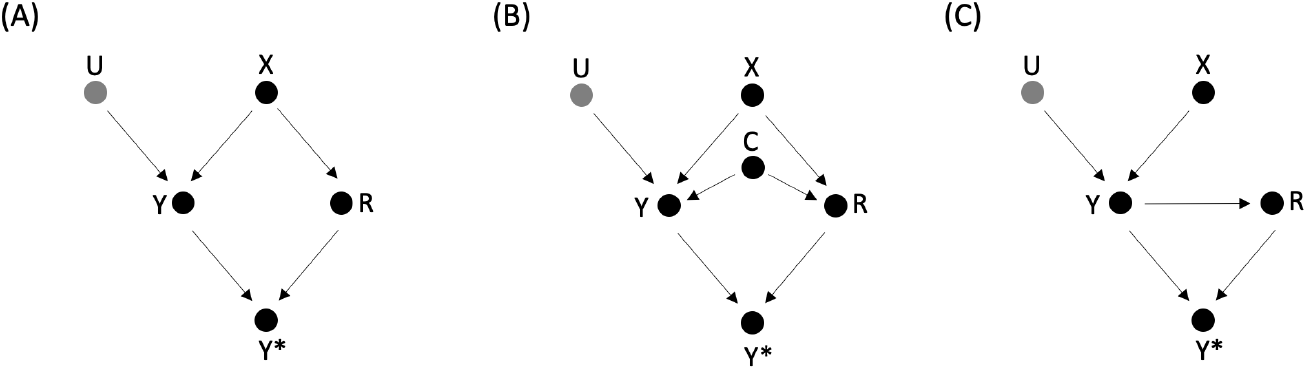
Three directed acyclic graphs (DAGs), depicting the relationship between binary treatment (*X*), continuous outcome (*Y*), an unmeasured continuous variable (*U*), a measured continuous variable (*C*) and the response indicator (*R*). The observed outcome is some function *f* (*Y, R*) and denoted *Y* ^*\**^. **A1)** Dropout dependent on *X*; **B)** Dropout dependent on *X* and *C*; **C)** Dropout dependent on *Y*.

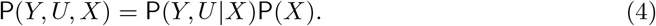

*X* d-separates *U* and *Y* from *R*, implying the conditional independence (*U, Y*) ╨ *R∣X*, so that

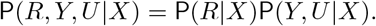

It follows that

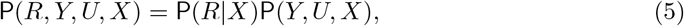

and rearranging (5) we obtain

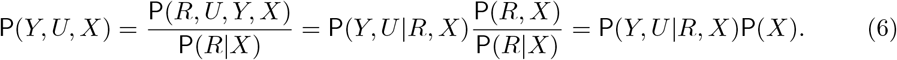

Then, from (4) and (6) we observe that the density of (*Y, U*) conditional on (*X, R*) and conditional on only *X* are the same:

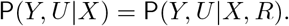

As P(*Y, U|X, R*) = P(*Y, U|X, R* = 1), with the latter the density of the observed outcome, (*Y* ^*\**^, *U*), we conclude that

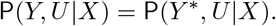

Then, for *X* = {0, 1}, *µ*_*X,R*=0_ = *µ*_*X,R*=1_ and 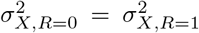. Under the additional assumption of homogeneous treatment effects, 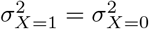, from which it follows that the variances in the observed sample are equal: 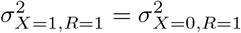

Similarly, for the MAR dropout mechanism in Figure 2.B, where *Y* is some function *f* (*X, A, U*), and *R* some function *g*(*X, C*), we obtain:

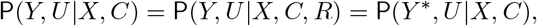

implying that the variance of the treatment groups in the observed sample, conditional on the observed variable *C*, are equal.

Figure 2.C depicts an MNAR dropout mechanism, with *Y* some function *f* (*X, U*) and *R* some function *g*(*Y*). *Y* d-separates *U* and *X* from *R*, implying the conditional independence (*U, X*) **╨** *R*|*Y*, so that

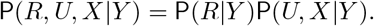

This can be rewritten as

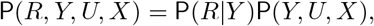

and

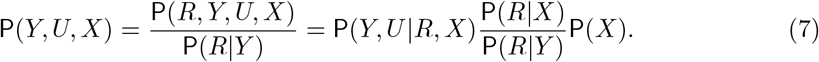

From (4) and (7) we note that

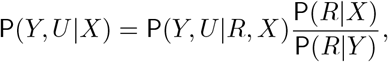

and that, under an MNAR dropout mechanism, the densities of (*Y, U*) conditional on (*X, R*) and conditional on *X* are no longer the same: P(*Y, U* |*X*) ≠ P(*Y, U* |*R, X*). If we assume treatment effect homogeneity (so that 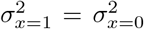), this difference between full and observed densities implies that the variances in the observed sample can be different. As MAR dropout results in no such difference, unequal observed group variances imply an MNAR dropout mechanism, which may, in turn indicate a biased estimate of the treatment effect. Note, however, that MNAR dropout does not necessarily result in a variance difference.

## 4 Methods for testing differences in variances

Various methods are available for testing and estimating the difference in variance between two groups, including Bartlett’s test ^22^, Levene’s test ^23^, the Brown-Forsythe test, ^24^, the Breusch-Pagan test ^25^, and the studentized Breusch-Pagan test ^26^. In this paper we employ the latter, as it has a straightforward implementation that allows for conditioning on additional covariates, and is also robust against non-normally distributed errors. This is particularly relevant, as, in practice, outcomes are unlikely to be strictly normally distributed. Even if the study sample is drawn from a population with normally distributed outcomes, the observed outcome distribution would, after non-random dropout, no longer be normal.

The studentized Breusch-Pagan estimate is obtained as follows. First, the outcome, *Y*, is regressed on the treatment variable, *X*, and optional additional covariates, *C*, in an OLS regression:

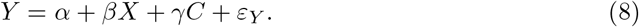

The regression residuals are obtained and squared 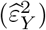 and, in a second auxiliary OLS regression, regressed on the treatment variable:

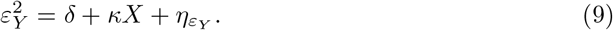

The variance difference estimate is given by *κ*, and the test statistic is given by *nR*^2^, with *n* the sample size and *R*^2^ obtained from the second, auxiliary regression.

## 5 A Simulation study

In this section, we examine in simulation the relationship between the bias of the CCA treatment effect, estimated with OLS linear regression, and the variance difference after dropout, estimated with the studentized Breusch-Pagan test.

### 5.1 Methods

We performed a 1000-fold simulation at sample sizes of *N*1 = 500, *N*2 = 1000 and *N*3 = 10000, for data with normally and log-normally distributed outcomes, and some unmeasured variable, *U*, acting on the outcome. Outcomes were simulated with treatment group variances of 8, 27% overall dropout (simulated under a logit model), with treatment effects of *β* = 1 and *β* = 0. Five data-generating scenarios were considered, which are shown as DAGs A-E in Figure 3, defined in the same manner as in Figure 2 (Section 3), with, e.g., in DAG B, *Y* MAR conditional on *X*. A detailed description of the simulation framework, in accordance with ADEMP guidelines ^27^, is given in Appendix A.1.

**Figure 3:**
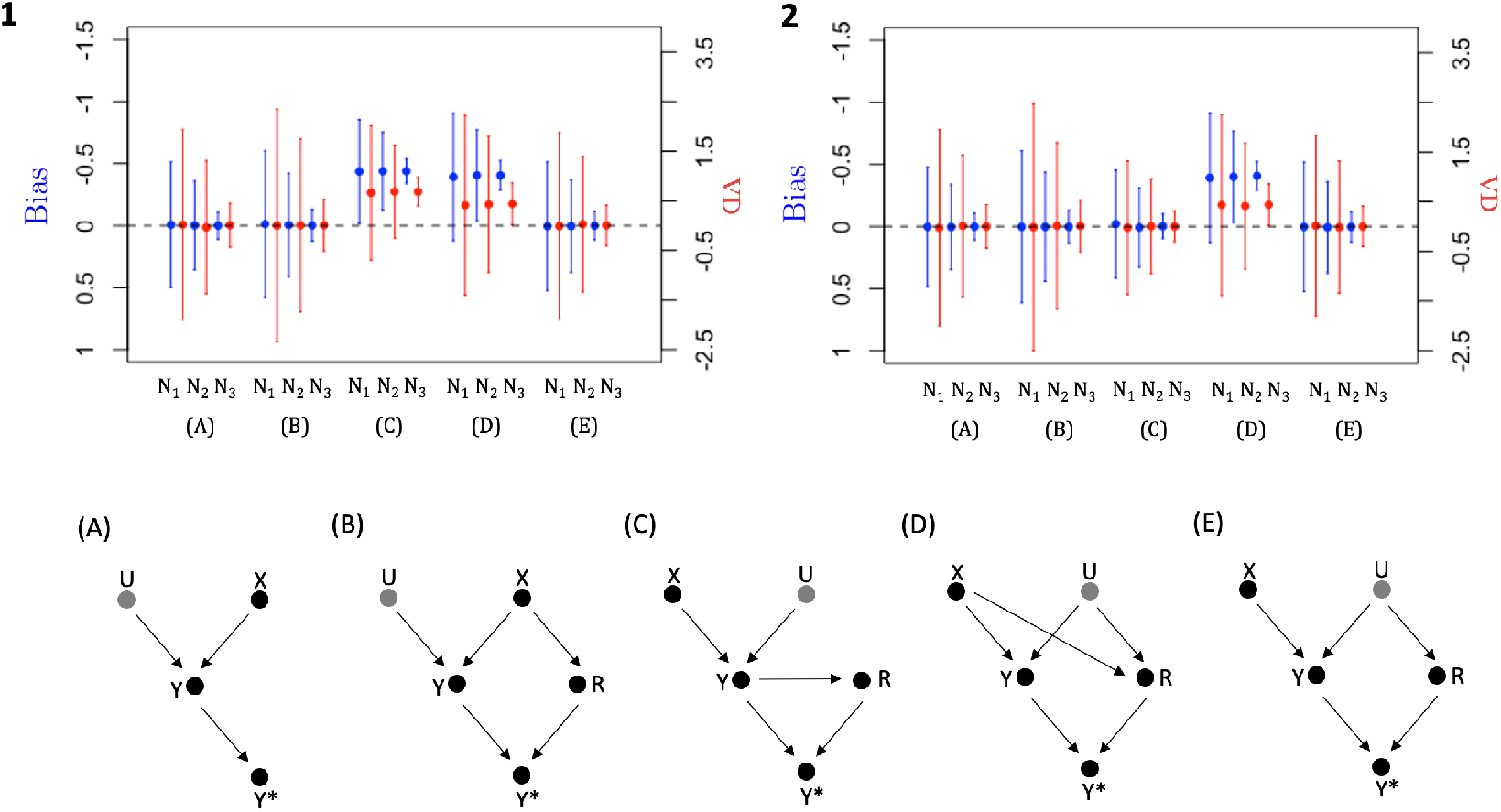
Bias of the complete case analysis (CCA) treatment effect (blue) on the left-hand Y-axis and the variance difference (VD, red) on the right-hand Y-axis in the observed sample with 95% confidence intervals (CIs), for data simulated according to DAGs A-E with a true treatment effect of (1) *β* = 1 and (2) *β* = 0. DAGs A-E represent **(A)** No dropout; **(B)** MAR dropout dependent on treatment; **(C)** MNAR dropout dependent on outcome, *Y*_*f*_ ; **(D)** MNAR dropout dependent on treatment and an unmeasured covariate, *U*, interacting on the probability scale; **(E)** MNAR dropout dependent on some unmeasured variable, *U*.

### 5.2 Results

Figure 3.1 summarizes the simulation results for a true treatment effect of *β* = 1, with on the left-hand Y-axis, in blue, the bias, and on the right-hand axis, in red, the variance difference after dropout, with a positive variance difference indicating a greater variance in the intervention group than the comparator group. In scenarios C and D, outcomes are MNAR and we observe bias and a positive variance difference.

In scenarios A, B, and E, the treatment effect estimate is unbiased with a zero variance difference. In scenarios A and B, the outcomes are fully observed and MAR, respectively. Even though *Y* is MNAR in scenario E (as *U* is unobserved), the treatment effect estimate is unbiased, as *U* and *X* are uncorrelated, and the estimate is obtained through OLS regression ^28^. Figure 3.2 shows the results under the null (*β* = 0), and we observe that all scenarios save D are unbiased, with corresponding zero variance differences. The power to detect a variance difference is smaller than the power to detect a treatment effect of the same size ^29^, and we observe that the confidence intervals (CIs) of the variance difference are wider than those of the bias. Figure A.1 (Appendix A.2) shows comparable results from simulations performed under different selection mechanism strengths. We observe similar results in Figure A.2, where the outcomes follow a log-normal distribution, but note that the CIs are wider than for normally distributed outcomes (Figure 3, Figure A.1). In Appendix A.3, companions tables to Figures A.1 and A.2 are provided for each DAG (Figure 3), with numerical estimates of the bias, variance difference, and additional quantities (Tables A.1-A.5 for DAGs A-E).

## 6 MNAR dropout and heterogeneous treatment effects in longitudinal data

In previous sections, we assumed homogeneous treatment effects, so that MNAR dropout was the only potential source of a variance difference in the observed outcome. Now, we consider longitudinal data, with outcomes measured at baseline and at final follow-up, and allow for the presence of effect modification (EM), which results in a heterogeneous treatment effect. We use ‘outcome at baseline’ to refer to a prognostic variable measured at baseline, which uses the same scale as the outcome measured at follow-up (e.g., in the applied example in Section 8.1, we consider the headache score measured at baseline and at 12 months). While EM will result in a variance difference at follow-up ^29^, it will not result in different treatment arm variances at baseline, in either the full or observed data. In contrast, outcome-dependent dropout may result in a variance difference at baseline in the observed data, as the outcome at baseline affects the outcome at follow-up, which in turn causes dropout. In the simulation below, we show that, for longitudinal data, the variance difference at baseline can be used to distinguish between treatment effect heterogeneity and outcome-dependent dropout.

### 6.1 Methods

We performed a 1000-fold simulation of *N* = 1000, with equal numbers of patients randomized to the intervention and comparator groups, a true treatment effect *β* = 1, 27% overall dropout (simulated under a logit model) and some unmeasured variable, *U*, acting on the outcome at final follow-up. Outcomes at baseline (*Y*_*b*_) and final follow-up (*Y*_*f*_) were drawn from a multivariate normal distribution, with variances of 4 and 5, respectively, and a correlation coefficient of 0.65. The variance difference of the observed sample was calculated at final follow-up (VD_*f*_) and, in the same set of patients, at baseline (VD_*b*_). CCA estimator bias and VD estimates were obtained with and without adjusting for *Y*_*b*_ (VD_*f*(*b*)_). The presence of treatment effect heterogeneity was assessed by testing if severity at baseline acts as an effect modifier. This was done by calculating the VD estimate adjusted for *Y*_*b*_ and the interaction between *Y*_*b*_ and treatment, *X* (VD_*f*(*bI*)_).

We considered four scenarios, analogous to scenarios A-D described in Section 5, but now with outcomes measured at baseline and at follow-up, and each without and with EM (4): A) No dropout; B) MAR dropout dependent on treatment, *X*; C) *Y*_*f*_ -dependent MNAR dropout D) MNAR dropout dependent on *X* and some unmeasured continuous covariate *U*, with *X* and *U* interacting on the log probability scale. In scenarios A(EM)-D(EM), *Y*_*b*_ acts as a positive effect modifier (i.e., patients with higher baseline values have a better treatment response and vice versa), simulated such that in the full data, EM increased the intervention group variance at final follow-up by 4. A detailed description of the simulation framework, in accordance with ADEMP guidelines ^27^, is given in Appendix B.1.

### 6.2 Results

The simulation results are shown in Table 1. We first consider the four scenarios when treatment effects are homogeneous (A-D). When dropout is absent (A) and when dropout is MAR conditional on treatment (B), the treatment effect estimates are unbiased, irrespective of conditioning on *Y*_*b*_, with on average zero variance differences at baseline (VD_*b*_) and at follow-up (VD_*f*_). When dropout is MNAR dependent on *Y*_*f*_ (C), we observe a biased treatment effect estimate and non-zero variance differences at baseline and at follow-up. Conditioning on *Y*_*b*_ results in an attenuated bias estimate and a smaller variance difference at follow-up (VD_*f*(*b*)_). In contrast, when dropout is MNAR independent of *Y*_*f*_ (D), we observe that there is no variance difference at baseline, and that conditioning on *Y*_*b*_ does not affect the bias estimate nor the variance difference at follow-up. For all scenarios, there is no effect of additionally conditioning on the *Y*_*b*_ and treatment interaction term when estimating the variance difference (VD_*f*(*bI*)_).

**Table 1:**
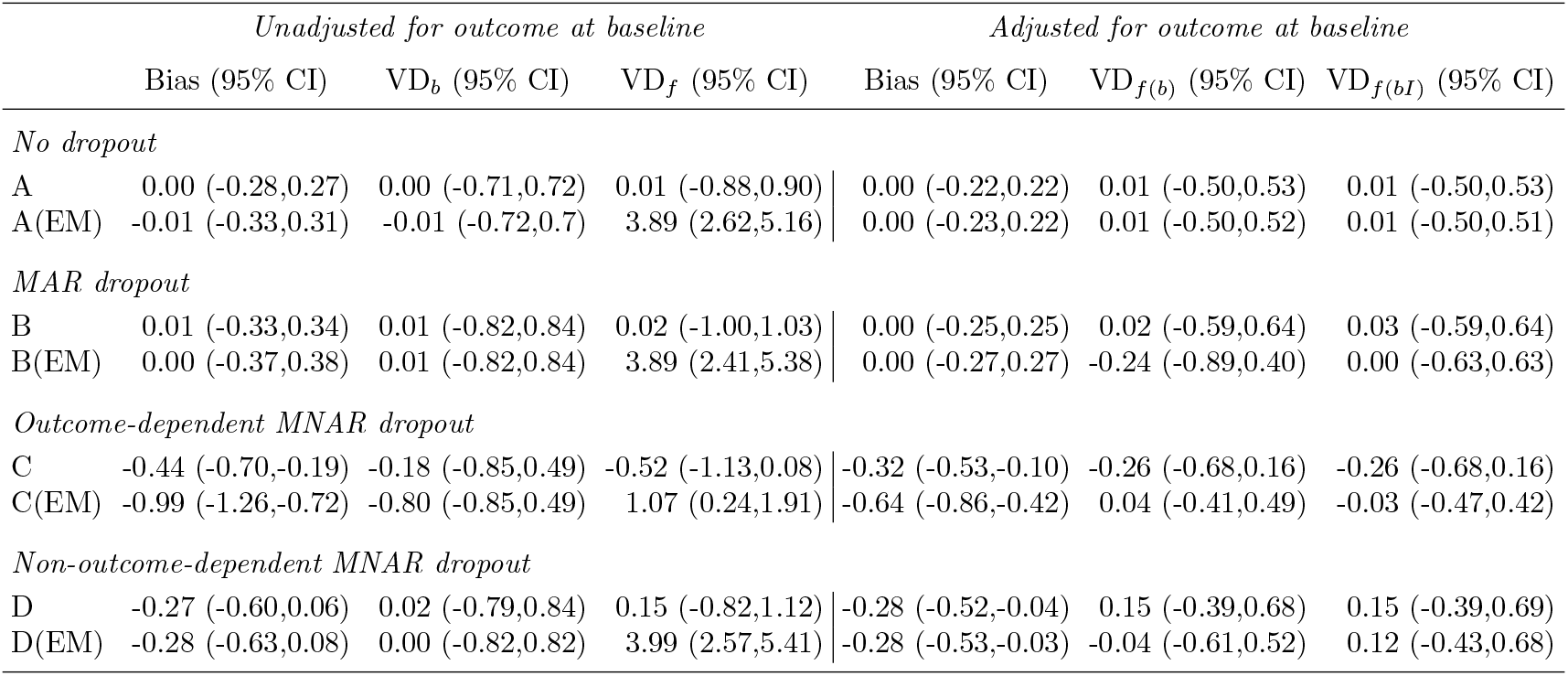
Bias of the complete case analysis (CCA) treatment effect estimate and variance difference in the observed sample with 95% confidence intervals (CIs), for longitudinal data, with baseline (*Y*_*b*_) and follow-up (*Y*_*f*_) measurements, simulated under different dropout mechanisms, without (A) and with (B) effect modification (EM). Of 1000 patients, 50% were randomized to each treatment group. Estimates were obtained without and with conditioning on the baseline outcome, with variance differences calculated at baseline (VD_*b*_) and at final follow-up, for the set of patients still observed at final follow-up. The variance difference at final follow-up was also calculated conditional on *Y*_*b*_ (VD_*f*(*b*)_), and conditional on *Y*_*b*_ and the interaction between *Y*_*b*_ and treatment (VD_*f*(*bI*)_). A) No dropout in both groups; B) MAR dropout dependent on treatment; C) MNAR dropout dependent on outcome, *Y*_*f*_ ; D) MNAR dropout dependent on treatment and an unmeasured covariate, *U*, interacting on the probability scale.

When *Y*_*b*_ acts as an effect modifier, resulting in treatment effect heterogeneity, we observe a variance difference at follow-up, regardless of dropout mechanism. In all four scenarios, conditioning on *Y*_*b*_ when estimating the variance difference (VD_*f*(*b*)_) results in a smaller variance difference, which is near zero in scenarios A, C and D. Additionally conditioning on the *Y*_*b*_ and treatment interaction term has little effect on scenarios A and C, but results in an increased variance difference in scenario D, and a zero variance difference in B. Table B.2 (Appendix B.3) describes the simulation results when unequal numbers of patients are randomized to treatment groups. There, we observe that when *Y*_*b*_ acts as an effect modifier, conditioning on *Y*_*b*_ results in a decreased VD_*f*(*b*)_, while additionally conditioning on the *Y*_*b*_ and treatment interaction term consistently results in a smaller VD_*f*(*bI*)_, regardless of dropout mechanism. In Appendix B.2 and B.3, companion tables are provided for Table 1 and Table B.2 (Table B.1 and Table B.3, respectively), which include measures of simulation performance.

In summary, a variance difference at baseline indicates outcome-dependent MNAR dropout, while a variance difference at follow-up may be the result of either outcome-dependent MNAR dropout, non-outcome-dependent MNAR dropout or heterogeneous treatment effects. A change in treatment effect estimate after conditioning on *Y*_*b*_ suggests either treatment effect heterogeneity, with baseline outcome affecting treatment response, or outcome-dependent MNAR dropout. A decrease in variance difference at follow-up, after conditioning on a variable, implies this variable either affects dropout or contributes to treatment effect heterogeneity. Specifically, conditioning on outcome at baseline, *Y*_*b*_, will decrease variance difference at follow-up when dropout is outcome-dependent or when *Y*_*b*_ is an effect modifier. We note that the size of the decrease in bias and variance difference depends on the correlation strength between *Y*_*b*_ and *Y*_*f*_, and, if present, the strength of EM, respectively. A further change in variance difference at follow-up when conditioning on *Y*_*b*_ and the interaction between *Y*_*b*_ and treatment implies treatment effect heterogeneity resulting from EM caused by *Y*_*b*_. When patients are randomized to unequally sized treatment groups, additionally conditioning on the *Y*_*b*_ and treatment interaction term consistently results in a decreased variance difference, when *Y*_*b*_ acts as an effect modifier.

## 7 Using conditional group variances to evaluate imputation models

In Section 3, we showed that for an MAR dropout mechanism, the group variances in the observed sample are equal, when conditioning on all variables that affect missingness. This property, in conjunction with the assumption of homogeneous treatment effects, can be used to assess the possibility of bias in an CCA analysis, by comparing the group variances while conditioning on all model variables. In an MI model, given that dropout is MAR conditional on the imputation model variables and the imputation model is correctly specified, we would expect the variance difference to be zero across the imputed datasets.

### 7.1 Methods

We performed a 1000-fold simulation of *N* = 1000 and *N* = 10000, a true treatment effect *β* = 1, 27% overall dropout (simulated under a logit model), variables *A* and *B* acting on outcome *Y*, for data with normally distributed outcomes and treatment group variances of 8. Two data-generating scenarios were considered, shown as DAGs in Figure 5, defined in the same manner as in Figure 2, with, e.g., in DAG M1, *Y* MAR conditional on *X* and *B*. We performed a CCA regression conditional on *A*, and an MI regression using the same analysis model, but with *B* additionally included in the imputation model. The corresponding variance differences for both models were estimated conditional on *A*. A detailed description of the simulation framework, in accordance with ADEMP guidelines ^27^, is given in Appendix C.1.

**Figure 4:**
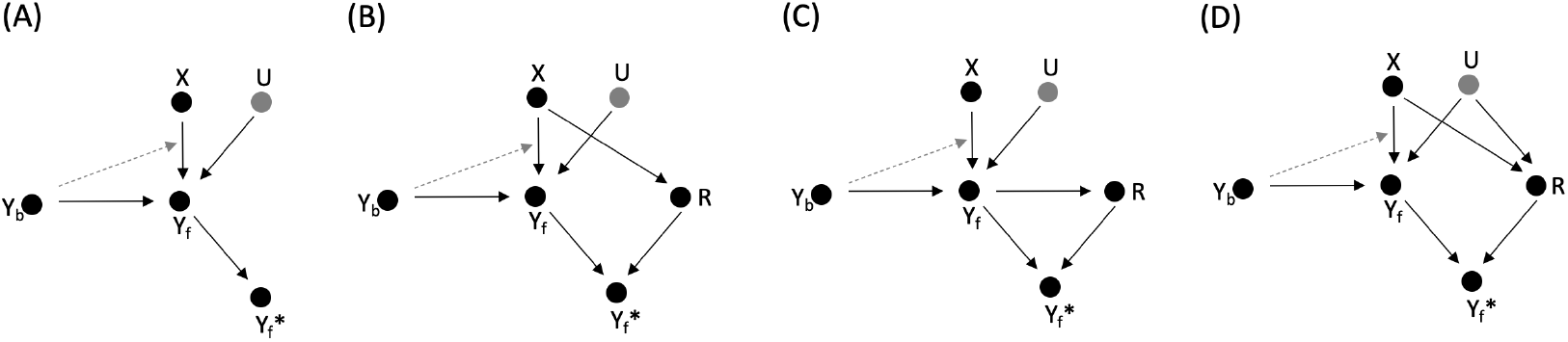
Four directed acyclic graphs (DAGs), depicting the relationship between binary treatment (*X*), continuous outcome at baseline (*Y*_*b*_), continuous outcome at final follow-up (*Y*_*f*_), unmeasured continuous variable (*U*), and the selection indicator (*R*). The observed outcome is some function *f* (*Y*_*f*_, *R*) and denoted 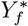. These four DAGs illustrate the data generating mechanisms used in the simulations of Table 1. Each scenario was considered with and without treatment effect heterogeneity, resulting from *Y*_*b*_ acting as an effect modifier, shown here as as gray dashed arrow. **(A)** No dropout; **(B)** MAR dropout dependent on treatment, *X*; **(C)** MNAR dropout dependent on *Y*_*f*_ ;**(D)** MNAR dropout dependent on *X* and *U*, which interact on the probability scale

**Figure 5:**
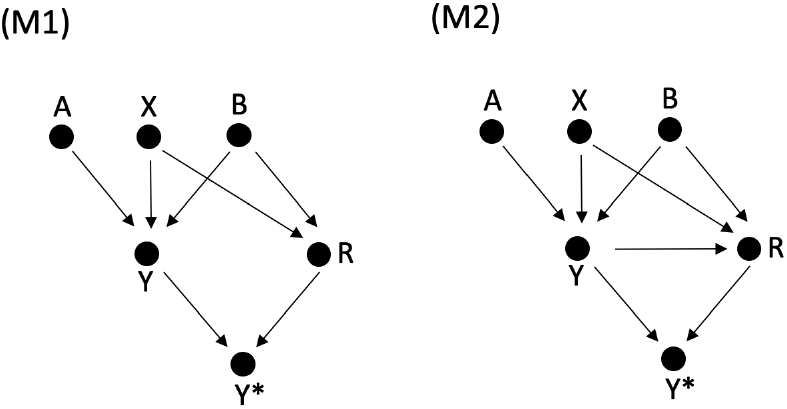
Two directed acyclic graphs (DAGs), depicting the relationship between binary treatment (*X*), continuous outcome (*Y*), two measured continuous variable (*A* and *B*) and the selection indicator (*R*). The observed outcome is some function *f* (*Y, R*) and denoted *Y* ^*\**^. **(M1)** *Y* is MAR conditional on *X* and *B*, with *X* and *B* interacting on the probability scale; **(M2)** *Y* MNAR, conditional on *X* and *B*, with dropout dependent on *X, B*, and *Y*, with *X* and *B* interacting on the log probability scale.

### 7.2 Results

The simulation results for *N* = 1000 are shown in Table 2. In scenario M1 (Figure 5.M1) dropout is MAR conditional on *X* and *B* and the CCA treatment effect estimate, obtained conditional on *A*, is biased, with the corresponding variance difference non-zero. Fitting the same analysis model, which regresses *Y* on *X* and *A*, to data imputed conditional on *Y, X, A* and *B*, results in an unbiased treatment effect estimate and a near-zero variance difference. In scenario M2 (Figure 5.M2), dropout is a function of *Y*, in addition to *B* and *X*, and the MI estimate is no longer unbiased and the variance difference remains positive, as the imputation model fails account for the effect of *Y* on dropout.

**Table 2:**
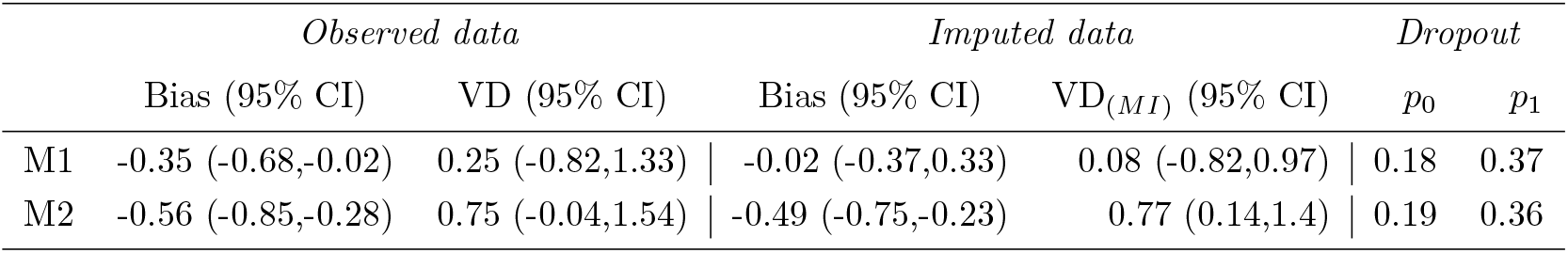
Bias of CCA and MI treatment effect estimates and variance differences (VDs) with 95% confidence intervals (CIs), for data (*N* = 1000) simulated according to DAGs M1 and M2 (Figure 5). M1) *Y* is a function of *A, B* and treatment, *X*, with dropout dependent on *A* and *B*; M2) Analogous to M1, with dropout additionally dependent on *Y*. Shown is the CCA treatment effect estimate, conditional on *A*, with corresponding VD, alongside the MI treatment effect estimate and VD (VD_*MI*_), estimated conditional on *A*, with both *A* and *B* included in the imputation model. Dropout proportions in the comparator and intervention group are denoted *p*_0_ and *p*_1_, respectively.

**Table 3:**
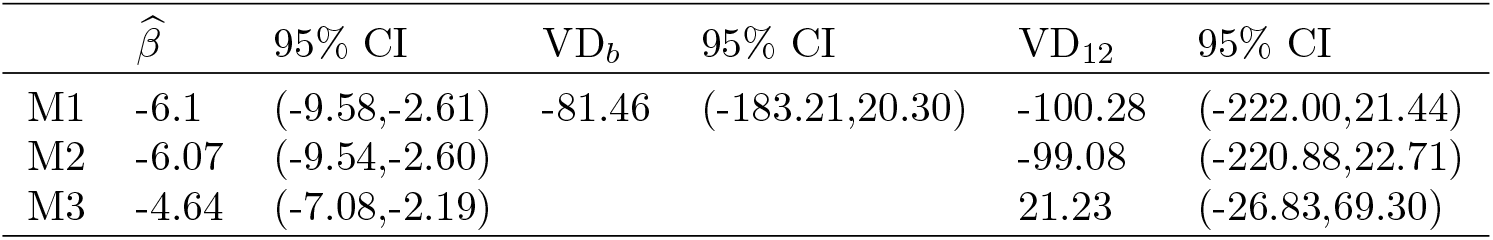
CCA treatment effect estimates and variance differences (VDs) with 95% confidence intervals (CIs) for data from the acupuncture RCT, which compared the effect of a treatment policy using acupuncture versus usual care on headache scores at 12 month follow-up. VDs were calculated at baseline (VD_*b*_) and at 12 months (VD_12_), for the set of patients still observed at 12 months. M1) Headache score is regressed on treatment; M2) Regression is adjusted for five minimization variables; M3) Regression is adjusted for five minimization variables and the baseline headache score (analysis model used in the published trial results).

| | $\hat{\beta}$ | 95% CI | $VD_b$ | 95% CI | $VD_{12}$ | 95% CI |
| --- | --- | --- | --- | --- | --- | --- |
| M1 | -6.1 | (-9.58,-2.61) | -81.46 | (-183.21,20.30) | -100.28 | (-222.00,21.44) |
| M2 | -6.07 | (-9.54,-2.60) |  |  | -99.08 | (-220.88,22.71) |
| M3 | -4.64 | (-7.08,-2.19) |  |  | 21.23 | (-26.83,69.30) |

Based on the variance difference in the observed and imputed data, we would conclude, for scenario M1, that including variable *B* in the imputation model will result in a less biased estimate, while for M2 we would infer that the imputation model fails to address the dropout mechanism, suggesting that the data are MNAR given the variables in the imputation model. In Appendix C.2, a companion table is provided for Table 2 (Table C.1), which includes results for a sample size of *N* = 10000 and various measures of simulation performance.

## 8 Application

### 8.1 An application using individual-level data from the acupuncture trial

We now apply our method to individual-level data from an RCT, which compared the effect of two treatments on 401 patients suffering from chronic headaches. The primary outcome was the headache score at 12 months, with higher values indicating worse symptoms ^20^. Patients were randomly allocated to acupuncture intervention (N=205) or usual care (N=196). The trial found a beneficial effect of acupuncture treatment, with a mean difference in headache score of -4.6 (95% CI: -7.0, -2.2), adjusted for baseline headache score and minimization variables age, sex, headache type, number of years of headache disorder, and site (general practices in England and Wales). At 12 months, 21% and 29% of patients in the acupuncture and usual care group, respectively, had dropped out. The investigators noted that while dropouts were generally comparable across the two groups, their baseline headache score was on average higher.

Using the variance differences at 12 months (VD_12_) and at baseline (VD_*b*_), we assessed the risk of bias due to MNAR dropout for an unadjusted CCA model (M1), a model adjusted for the minimization variables (M2), and a model adjusted for the minimization variables and baseline headache score (M3). VD_12_ and VD_*b*_ were estimated using the studentized Breusch-Pagan test, for the subset of patients still observed at 12 months.

The unadjusted model (M1), regressing headache score on treatment, showed a beneficial effect of acupuncture therapy (−6.1, 95% CI:-9.6,-2.6) and lower outcome variances for the acupuncture group at baseline and at 12 months, with VD_*b*_=-81.5 (95% CI: -183.2,20.3) and VD_12_=-100.3 (95% CI: -222.0,21.4), respectively. Adjusting for the five minimization variables (M2), did not affect the estimated treatment effect or VD_12_, while additionally adjusting for baseline headache score resulted in an attenuated treatment effect of -4.64 (95% CI: -7.08,-2.19) and a near zero VD_12_, suggesting that baseline headache score may act as an effect modifier, with the observed VD_12_ the result of EM in the intervention group, or that dropout is MNAR dependent on outcome. The variances were also different at baseline (VD_*b*_), which points to outcome-dependent MNAR dropout. In summary, our results suggest that the CCA estimate, adjusted for minimization variables and baseline headache score may be biased due to outcome-dependent dropout, and that the true effect may be more modest. This bias is, however, likely partly adjusted for by conditioning on baseline headache score. A next step would involve performing an MNAR sensitivity analysis.

### 8.2 An application using summary-level data from the POPPI trial

The POPPI trial investigated whether a preventive, complex psychological intervention, initiated in the intensive care unit (ICU), would reduce the development of subsequent post-traumatic stress disorder (PTSD) symptoms at 6 months in ICU patients ^30^. Symptom severity was quantified using the PTSD Symptom scale-self-report (PSS-SR) questionnaire, with higher values indicating greater severity. Twenty-four ICUs were randomized to intervention or control, with intervention ICUs providing usual care during a baseline period and the preventive intervention during the intervention period, and control ICUs providing usual care throughout. At 6 months follow-up, 79.3% of patients had completed the PSS-SR questionnaire, with no difference across study arms. The trial found no beneficial effect of intervention, with a mean difference in PSS-SR score of -0.03 (95% CI: -2.58, 2.52), adjusted for age, sex, race/ethnicity, deprivation, preexisting anxiety/depression, planned admission following elective surgery and the Intensive Care National Audit Research Centre (ICNARC) Physiology Score.

Using summary statistics from the published study, we performed a t-test for the variance difference ^29^ between groups at 6 months, to assess if the study’s reported null result may have been biased by MNAR dropout. Published estimates were means with 95% CIs, adjusted for the previously listed variables, which we used to calculate the variances and corresponding VDs. We found no evidence for a variance difference across groups at baseline (VD_*b*_=11.2; 95% CI: -22.7,45.2), but a greater variance in the intervention group at 6 months (VD_6_=52.5; 95% CI: 18.8,86.2).

No variance difference across groups at baseline indicates dropout is not outcome-dependent, while the variance difference at follow-up suggests there may be treatment effect heterogeneity or MNAR dropout that does not depend on outcome. The latter, however, is unlikely, as dropout was balanced across treatment groups. For non-outcome-dependent dropout to result in bias and a variance difference, this requires an interaction with treatment in the dropout mechanism. This would have resulted in unbalanced dropout. In summary, our results suggest there is no MNAR dropout, but that there may be treatment effect heterogeneity, which, on average, results in a null treatment effect.

## Discussion

In this paper, we show that in RCTs, under the assumption of homogeneous treatment effects, a difference in observed outcome variances between treatment groups, conditional on the model variables, implies an MNAR dropout mechanism and by extension a biased CCA treatment effect estimate. In contrast, when outcomes are MAR, the conditional treatment group variances will be equal and the CCA estimate will be unbiased.

Treatment effect heterogeneity can be thought of as non-random variability in treatment response that is attributable to patient characteristics. How plausible it is that heterogeneity is absent depends strongly on intervention type and study population. Efficacy trials, for example, typically have stricter inclusion criteria, resulting in more homogeneous study populations, and are less prone to large variations in treatment response. In contrast, pragmatic trials with broad eligibility criteria are more likely to have heterogeneous treatment effects ^31^. As treatment effect heterogeneity affects the intervention group variance ^29^, it is a second potential cause of observed outcome variance differences. We show that in longitudinal trials, we can isolate the effect of outcome-dependent MNAR dropout by considering the variance difference at baseline. This baseline variable can be a baseline prognostic variable measured on the same scale as the final outcome, as in the applied examples in Section 8, or alternatively, any baseline prognostic variable that has a causal effect on the outcome of interest.

Dealing with missing outcomes is challenging, as the underlying dropout mechanism typically cannot be inferred from the observed data. Existing approaches for investigating whether data are MNAR involve examining covariate distributions and dropout proportions, and using expert knowledge and/or auxiliary information to assess the plausibility of outcome-related dropout ^10–12^. Despite the availability of useful methods and institutions like the NRC and EMA strongly encouraging improved practice ^13,14^, the vast majority of trials do not undertake or report any kind of sensitivity analysis or, indeed, perform any kind of risk of bias assessment ^15–19^.

We propose employing the (conditional) variance difference as a MNAR bias assessment tool, and, indirectly, as a model building tool, which can be used to assess the added value of including variables for explaining the missingness mechanism. This method is easily implemented, using existing tests available in standard software, and has a straightforward interpretation of results. In Section 8, we demonstrated how variance differences can be used to assess the risk of MNAR bias for various models, using both individual-level data from the acupuncture trial, and summary-level data from the POPPI trial.

## Data Availability

The individual-level patient data used in this study are available from
https://www.causeweb.org/tshs/acupuncture/.

