## Appendix for "Treatment group outcome variance difference after dropout as an indicator of missing-not-at-random bias in randomized clinical trials"

This document provides more detail on the simulation approach alongside additional results for the simulations reported in Sections 5, 6 and 7 of the main text, in appendices A, B and C, respectively.

#### Appendix A

This appendix gives a detailed description and the results of the simulation study of Section 5 of the main test in full. The bias of the treatment effect, as estimated by a CCA linear regression, and the variance difference, as estimated by the studentized Breusch-Pagan test, are investigated through simulations of five missing data mechanisms (see the DAGs of Figure 3 in the main text). Figure A.1 summarizes the results for normally distributed outcomes, Figure A.2 for log-normally distributed outcomes. Companion tables to Figure A.1 and A.2 are also provided for each of the DAGs (Figure 2 in the main text), with numerical estimates of bias, variance, and additional quantities (Tables A.1-A.5 for DAGs A-E). Section A.1 gives a detailed description of the simulation approach. Section A.2 shows the results in plots. Section A.3 provides the companion tables for each DAG.

##### A.1 Simulation approach

Here, we describe the simulation approach in detail using the ADEMP framework, and define the aims, data-generating mechanisms, estimands, methods, and performance measures.

**Aim**: Illustrating that 1) dropout that is MAR conditional on the model variables does not result in bias and no variance difference; 2) dropout that is MNAR conditional on the model variables results in bias and a variance difference

**General setup:** Data with normally and log-normally distributed outcomes, simulated with a treatment effect and under the null, at three different sample sizes, for five different dropout scenarios, simulated with a logit mechanism at two different strengths

Data generating mechanism:

**Variables**

1. Binary treatment variable, $X \sim B(1, 0.5)$, some normally distributed variable $U \sim N(0, 4)$, which affects Y, so that $Y = \beta X + U + \varepsilon$, with treatment effect $\beta$ and $\varepsilon \sim N(0, 4)$, so that $\text{var}(Y|X) = 8$. (Results reported in main text, Figure 1 and Supplementary Figure 1)
2. Binary treatment variable, $X \sim B(1, 0.5)$, some log-normally distributed variable $\log(U) \sim N(2.283, 0.198)$, zero-centered after simulation, which affects Y, so that $Y = \beta X + U + e$, with treatment effect $\beta$ and $\log(\varepsilon) \sim N(2.283, 0.198)$, zero-centered after simulation, so that $\text{var}(Y|X) = 8$. Parameters for the log-normal distribution were chosen such that the null hypothesis of the Shapiro-Wilkes test for normality was consistently rejected across simulated datasets (coverage of 0.938 for $N = 500$, 1 for $N = 1000$), and such that the mean standard error (SE) and the Monte Carlo SE (MCSE) were consistent with each other and comparable to the ones obtained when simulating normally distributed outcomes. (Results reported in Appendix, Tables A.1 to A.5 for dropout scenarios A to E)

**Treatment effect:** $\beta = 1$ and $\beta = 0$

**Sample size:** Total sample sizes of $N = 500$, $N = 1000$ and $N = 10000$.

**Dropout mechanism:** Logit dropout model, at two different strengths (coefficients of 1 and 2, for a 'weak' and 'strong' selection mechanism, respectively), with intercepts selected such that the overall dropout proportion was / 0.27, for five different mechanisms (shown as DAGs A to E in Figure 1 of the main text). For each mechanism, the probability of selection is defined conditional on the model variables: $\mathsf{P}(R = 1|Y, X, U)$, with $R$ the response indicator.

A *No dropout*
Normally and log-normally distributed $Y$ and $U$, for $\beta = 1$ and $\beta = 0$

- $\mathsf{P}(R = 1|Y, X, U) = 1$

B *$X$-dependent dropout*
Normally and log-normally distributed $Y$ and $U$, for $\beta = 1$ and $\beta = 0$

- $\mathsf{P}(R = 1|Y, X, U) = \exp(0.55 + X)/(1 + \exp(0.55 + X)$
- $\mathsf{P}(R = 1|Y, X, U) = \exp(0.25 + 2X)/(1 + \exp(0.25 + 2X)$

C *$Y$-dependent dropout*
Normally distributed $Y$ and $U$, for $\beta = 1$

- $\mathsf{P}(R = 1|Y, X, U) = \exp(1.6 + Y)/(1 + \exp(1.6 + Y)$
- $\mathsf{P}(R = 1|Y, X, U) = \exp(2.6 + 2Y)/(1 + \exp(2.6 + 2Y)$

Normally distributed $Y$ and $U$, for $\beta = 0$

- $\mathsf{P}(R = 1|Y, X, U) = \exp(2 + Y)/(1 + \exp(2 + Y)$

- $\mathsf{P}(R=1|Y,X,U) = \exp(3.6+2Y)/(1+\exp(3.6+2Y)$

Log-normally distributed $Y$ and $U$, for $\beta = 1$

- $\mathsf{P}(R=1|Y,X,U) = \exp(1.6+Y)/(1+\exp(1.6+Y)$
- $\mathsf{P}(R=1|Y,X,U) = \exp(2.8+2Y)/(1+\exp(2.8+2Y)$

Log-normally distributed $Y$ and $U$, for $\beta = 0$

- $\mathsf{P}(R=1|Y,X,U) = \exp(2.1+Y)/(1+\exp(2.1+Y)$
- $\mathsf{P}(R=1|Y,X,U) = \exp(3.8+2Y)/(1+\exp(3.8+2Y)$

D *Non-$Y$-dependent MNAR dropout ($X$- and $U$-dependent*

Normally distributed $Y$ and $U$, for $\beta = 1$ and $\beta = 0$

- $\mathsf{P}(R=1|Y,X,U) = \exp(1.15+X+U)/(1+\exp(1.15+X+U)$
- $\mathsf{P}(R=1|Y,X,U) = \exp(1.7+2X+2U)/(1+\exp(1.7+2X+2U)$

Log-normally distributed $Y$ and $U$, for $\beta = 1$ and $\beta = 0$

- $\mathsf{P}(R=1|Y,X,U) = \exp(1.2+X+U)/(1+\exp(1.2+X+U)$
- $\mathsf{P}(R=1|Y,X,U) = \exp(1.9+2X+2U)/(1+\exp(1.9+2X+2U)$

E *$U$-dependent dropout*

Normally distributed $Y$ and $U$, for $\beta = 1$ and $\beta = 0$

- $\mathsf{P}(R=1|Y,X,U) = \exp(1.6+U)/(1+\exp(1.6+U)$
- $\mathsf{P}(R=1|Y,X,U) = \exp(2.7+2U)/(1+\exp(2.7+2U)$

Log-normally distributed $Y$ and $U$, for $\beta = 1$ and $\beta = 0$

- $\mathsf{P}(R=1|Y,X,U) = \exp(1.65+U)/(1+\exp(1.65+U)$
- $\mathsf{P}(R=1|Y,X,U) = \exp(2.8+2U)/(1+\exp(2.8+2U)$

**Simulation size:** For each scenario, 1000 datasets were simulated. Simulation quality was verified by checking of mean SEs of the CCA and variance difference estimates were comparable to the Monte Carlo SEs (see ‘Performance measures’)

**Estimands**

*Primary:* Reported in main text, Figure 3, and in appendix, Supplementary Figures 1 and 2.

- CCA treatment effect estimate, with 95% CI
- Variance difference across treatment groups after dropout, with 95% CI (with a positive variance difference indicating a greater variance in the treatment group than the comparator group)

*Secondary:* Reported in appendix, Tables A1 to A5, for dropout scenarios A to E.

- 95% coverage* of the two measures listed above
- Dropout proportion per group
- Standard error (SE) of the two measures listed above

- Monte Carlo SE (MCSE) of the two measures listed above
- Individual group means after dropout
- Group outcome variances after dropout

(* The 95% coverage refers to the proportion of times the estimate was excluded from the 95% CI. If the true estimate is 0, this should come to 0.05)

**Methods**

- CCA estimator
- Studentized Breusch-Pagan test (for estimating the variance difference across groups after dropout)
- 95% CIs were calculated using the Monte Carlo Standard error (MCSE)

**Performance measures**: Simulation quality was checked by seeing if the estimate SEs were comparable to the MCSEs

#### A.2 Results: figures

**Figure A.1:** Bias of the complete case analysis (CCA) treatment effect estimate and variance difference (VD) in the observed sample with 95% confidence intervals, for simulated ($S = 1000$) normally distributed outcome data, with treatment group variances of 8 ($\sigma_j^2 = 8$)), and 27% overall dropout. Data were simulated according to DAGs A-E in Figure 3 of the main text, for three sample sizes ($N1 = 500$, $N2 = 1000$ and $N3 = 10000$), with a treatment effect of $\beta = 1$ (**1**) and under the null (**2**), for a strong dropout mechanism, and with $\beta = 1$ (**3**) and under the null (**4**), for a weak dropout mechanism.

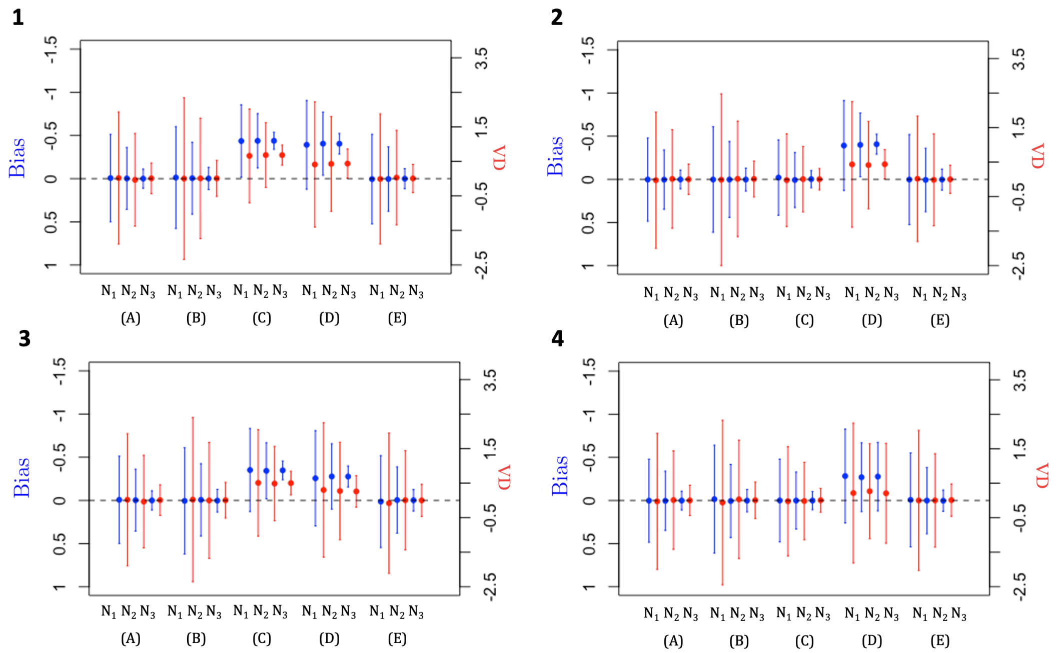

**Figure A.2:** Bias of the complete case analysis (CCA) treatment effect estimate and variance difference (VD) in the observed sample with 95% confidence intervals, for simulated ($S = 1000$) log-normally distributed outcome data, with treatment group variances of 8 ($\sigma_j^2 = 8$)), and 27% overall dropout. Data were simulated according to DAGs A-E in Figure 3 of the main text, for three sample sizes ($N1 = 500$, $N2 = 1000$ and $N3 = 10000$), with a treatment effect of $\beta = 1$ (**1**) and under the null (**2**), for a strong dropout mechanism, and with $\beta = 1$ (**3**) and under the null (**4**), for a weak dropout mechanism.

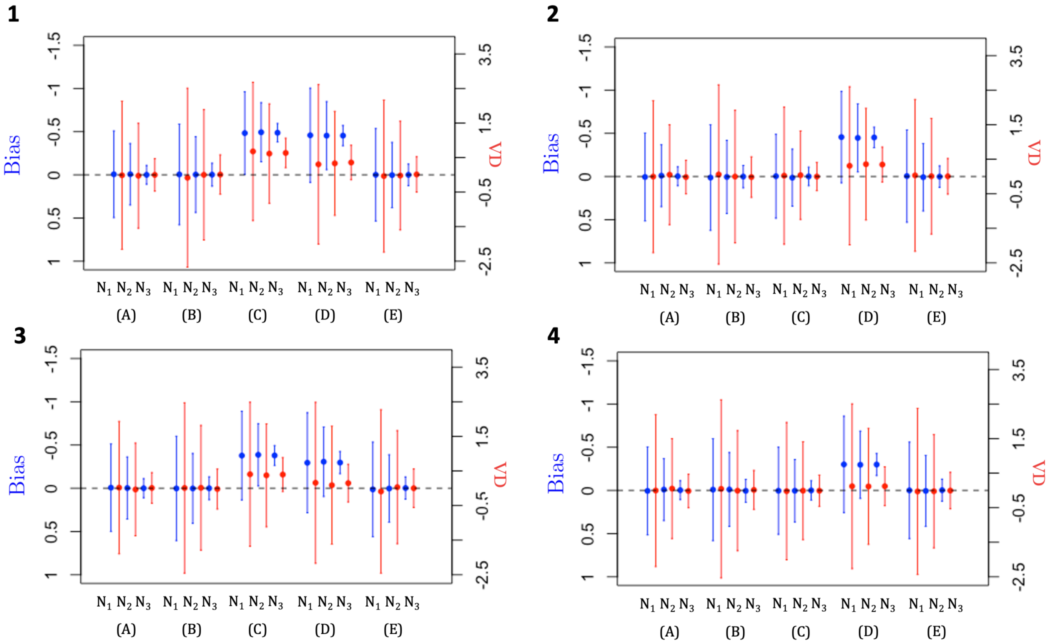

#### A.3 Results: companion tables

**Table A.1:** Bias of the treatment effect and variances for data simulated ($S = 1000$) with no dropout, according to DAG A. Data are simulated $S = 1000$ with intervention group mean and variance $\mu_1 = 1$ and $\sigma_1^2 = 8$), comparator group mean and variance $\mu_0 = 0$ and $\sigma_0^2 = 8$), for sample sizes of $N1 = 500$, $N2 = 1000$ and $N3 = 10000$. We distinguish between data with normally distributed errors for $Y$ and $U$, and log-normally distributed errors. **1)** Treatment effect of $\beta = 1$. **2)** Under the null ($\beta = 0$). Shown are the bias and variance difference (VD), with 95% confidence confidence interval (CI), standard error (SE), Monte Carlo SE (MCSE), and proportion of 95% CIs excluding the null ($p_{95\%\mathrm{CI}}$). Also included are the group means ( $\widehat{\mu}_1^*$, $\widehat{\mu}_0^*$) and the group variances ($\widehat{\sigma}_0^{2*}$, $\widehat{\sigma}_1^{2*}$).

| | Size | Bias | 95% CI | SE | MCSE | $p_{95\%\mathrm{CI}}$ | VD | 95% CI | SE | MCSE | $p_{95\%\mathrm{CI}}$ | $\widehat{\mu}_1^*$ | $\widehat{\mu}_0^*$ | $\widehat{s}_1^{2*}$ | $\widehat{s}_0^{2*}$ |
|---|---|---|---|---|---|---|---|---|---|---|---|---|---|---|---|
| | | | | | | | Normally distributed errors | | | | | | | | |
| | N1 | -0.01 | (-0.51,0.50) | 0.25 | 0.26 | 0.055 | 0.02 | (-1.89,1.93) | 1.01 | 0.98 | 0.045 | 1.00 | 0.00 | 8.02 | 8 |
| 1 | N2 | 0.00 | (-0.36,0.36) | 0.18 | 0.18 | 0.051 | -0.04 | (-1.37,1.30) | 0.71 | 0.68 | 0.043 | 1.00 | 0.00 | 7.99 | 8.03 |
| | N3 | 0.00 | (-0.11,0.11) | 0.06 | 0.06 | 0.053 | 0.01 | (-0.43,0.45) | 0.23 | 0.23 | 0.045 | 1.00 | 0.00 | 8.01 | 8 |
| | N1 | 0.01 | (-0.50,0.51) | 0.25 | 0.26 | 0.056 | 0.00 | (-2.20,2.20) | 1.08 | 1.12 | 0.056 | 0.00 | 0.00 | 7.98 | 7.99 |
| 2 | N2 | -0.01 | (-0.37,0.35) | 0.18 | 0.18 | 0.057 | 0.05 | (-1.39,1.50) | 0.77 | 0.74 | 0.039 | 0.00 | 0.00 | 8.01 | 7.96 |
| | N3 | 0.00 | (-0.11,0.11) | 0.06 | 0.06 | 0.05 | -0.02 | (-0.50,0.47) | 0.24 | 0.25 | 0.05 | 0.00 | 0.00 | 7.99 | 8.01 |
| | | | | | | | Log-normally distributed errors | | | | | | | | |
| | N1 | -0.01 | (-0.50,0.48) | 0.25 | 0.25 | 0.047 | -0.02 | (-2.11,2.07) | 1.08 | 1.07 | 0.043 | 0.99 | 0.01 | 7.98 | 8 |
| 1 | N2 | 0.01 | (-0.34,0.35) | 0.18 | 0.18 | 0.053 | -0.02 | (-1.49,1.45) | 0.77 | 0.75 | 0.043 | 1.00 | 0.00 | 7.99 | 8.01 |
| | N3 | 0.00 | (-0.11,0.11) | 0.06 | 0.05 | 0.047 | 0.00 | (-0.47,0.48) | 0.24 | 0.24 | 0.045 | 1.00 | 0.00 | 8.00 | 7.99 |
| | N1 | 0.00 | (-0.50,0.50) | 0.25 | 0.26 | 0.052 | -0.03 | (-2.15,2.09) | 1.08 | 1.08 | 0.05 | 0.00 | 0.00 | 8.00 | 8.03 |
| 2 | N2 | -0.01 | (-0.36,0.34) | 0.18 | 0.18 | 0.048 | -0.02 | (-1.53,1.50) | 0.77 | 0.77 | 0.045 | -0.01 | 0.01 | 8.00 | 8.02 |
| | N3 | 0.00 | (-0.12,0.12) | 0.06 | 0.06 | 0.064 | 0.00 | (-0.49,0.49) | 0.24 | 0.25 | 0.058 | 0.00 | 0.00 | 8.00 | 8 |

**Table A.2:** Bias of the treatment effect and variances after dropout for data simulated ($S = 1000$) under a treatment-dependent MAR dropout mechanism according to DAG B. Data are simulated $S = 1000$ with intervention group mean and variance $\mu_1 = 1$ and $\sigma_1^2 = 8$), comparator group mean and variance $\mu_0 = 0$ and $\sigma_0^2 = 8$), 27% overall dropout, for sample sizes of $N1 = 500$, $N2 = 1000$ and $N3 = 10000$. We distinguish between data with normally distributed errors for $Y$ and $U$, and log-normally distributed errors. **1)** Strong logit dropout mechanism in the presence of a treatment effect ($\beta = 1$). **2)** Strong logit dropout mechanism under the null ($\beta = 0$). **3)** Weak logit dropout mechanism in the presence of a treatment effect ($\beta = 1$). **4)** Weak logit dropout mechanism under the null ($\beta = 0$). Shown are the bias and variance difference (VD), with 95% confidence confidence interval (CI), standard error (SE), Monte Carlo SE (MCSE), and proportion of 95% CIs excluding the null ($p_{95\%\text{CI}}$). Also included are the group means after dropout ( $\widehat{\mu}_1^*$, $\widehat{\mu}_0^*$), the group variances after dropout ($\widehat{\sigma}_0^{2*}$, $\widehat{\sigma}_1^{2*}$), and the dropout proportions per treatment group ($p_0$,$p_1$).

| | Size | Bias | 95% CI | SE | MCSE | $p_{95\%\text{CI}}$ | VD | 95% CI | SE | MCSE | $p_{95\%\text{CI}}$ | $\widehat{\mu}_1^*$ | $\widehat{\mu}_0^*$ | $\widehat{s}_1^{2*}$ | $\widehat{s}_0^{2*}$ | $p_1$ | $p_0$ |
|---|---|---|---|---|---|---|---|---|---|---|---|---|---|---|---|---|---|
| | | | | | | | Normally distributed errors | | | | | | | | | | |
| | N1 | -0.01 | (-0.60,0.58) | 0.30 | 0.3 | 0.048 | 0.00 | (-2.34,2.34) | 1.20 | 1.19 | 0.041 | 1.00 | 0.02 | 8.00 | 8.02 | 0.09 | 0.44 |
| 1 | N2 | -0.01 | (-0.42,0.41) | 0.22 | 0.21 | 0.044 | 0.01 | (-1.73,1.75) | 0.86 | 0.89 | 0.049 | 1.01 | 0.01 | 8.01 | 8.01 | 0.10 | 0.44 |
| | N3 | 0.00 | (-0.13,0.13) | 0.07 | 0.07 | 0.043 | 0.01 | (-0.51,0.53) | 0.27 | 0.26 | 0.042 | 1.00 | 0.00 | 7.99 | 7.99 | 0.10 | 0.44 |
| | N1 | 0.01 | (-0.60,0.62) | 0.30 | 0.31 | 0.054 | 0.06 | (-2.53,2.65) | 1.29 | 1.32 | 0.044 | 0.00 | -0.01 | 8.01 | 7.97 | 0.09 | 0.44 |
| 2 | N2 | 0.00 | (-0.42,0.43) | 0.22 | 0.22 | 0.052 | 0.00 | (-1.92,1.92) | 0.92 | 0.98 | 0.062 | 0.00 | 0.00 | 8.01 | 8.02 | 0.10 | 0.44 |
| | N3 | 0.00 | (-0.13,0.13) | 0.07 | 0.07 | 0.042 | -0.02 | (-0.60,0.57) | 0.29 | 0.30 | 0.053 | 0.00 | 0.00 | 8.00 | 8.01 | 0.10 | 0.44 |
| | N1 | -0.01 | (-0.62,0.60) | 0.30 | 0.31 | 0.056 | 0.03 | (-2.31,2.37) | 1.18 | 1.19 | 0.053 | 0.99 | 0.00 | 8.01 | 7.99 | 0.18 | 0.36 |
| 3 | N2 | 0.00 | (-0.42,0.43) | 0.21 | 0.22 | 0.051 | -0.01 | (-1.68,1.65) | 0.84 | 0.85 | 0.048 | 0.99 | -0.01 | 7.97 | 7.99 | 0.17 | 0.37 |
| | N3 | 0.00 | (-0.13,0.13) | 0.07 | 0.07 | 0.05 | 0.00 | (-0.53,0.53) | 0.27 | 0.27 | 0.05 | 1.00 | 0.00 | 8.00 | 8 | 0.18 | 0.37 |
| | N1 | 0.00 | (-0.59,0.58) | 0.30 | 0.3 | 0.048 | 0.00 | (-2.29,2.30) | 1.18 | 1.17 | 0.051 | 0.00 | 0.00 | 7.98 | 7.99 | 0.18 | 0.37 |
| 4 | N2 | -0.01 | (-0.43,0.42) | 0.21 | 0.22 | 0.053 | 0.02 | (-1.61,1.66) | 0.84 | 0.84 | 0.044 | 0.00 | 0.00 | 7.98 | 7.96 | 0.18 | 0.37 |
| | N3 | 0.00 | (-0.13,0.13) | 0.07 | 0.07 | 0.046 | 0.00 | (-0.52,0.53) | 0.27 | 0.27 | 0.048 | 0.00 | 0.00 | 8.00 | 8 | 0.18 | 0.37 |
| | | | | | | | Log-normally distributed errors | | | | | | | | | | |
| | N1 | 0.00 | (-0.59,0.60) | 0.30 | 0.3 | 0.047 | 0.03 | (-2.44,2.50) | 1.29 | 1.26 | 0.046 | 1.00 | 0.00 | 7.99 | 7.99 | 0.10 | 0.44 |
| 1 | N2 | 0.00 | (-0.42,0.43) | 0.21 | 0.22 | 0.061 | 0.02 | (-1.83,1.87) | 0.92 | 0.94 | 0.054 | 1.00 | 0.00 | 8.00 | 8 | 0.10 | 0.44 |
| | N3 | 0.00 | (-0.13,0.13) | 0.07 | 0.07 | 0.044 | 0.00 | (-0.56,0.55) | 0.29 | 0.28 | 0.043 | 1.00 | 0.00 | 8.01 | 8.01 | 0.09 | 0.44 |
| | N1 | -0.01 | (-0.58,0.56) | 0.30 | 0.29 | 0.04 | 0.04 | (-2.46,2.54) | 1.29 | 1.28 | 0.041 | 0.00 | 0.01 | 8.03 | 8.01 | 0.10 | 0.44 |
| 2 | N2 | 0.00 | (-0.43,0.42) | 0.21 | 0.22 | 0.053 | -0.01 | (-1.83,1.81) | 0.92 | 0.93 | 0.058 | 0.00 | 0.00 | 7.99 | 8.01 | 0.10 | 0.44 |
| | N3 | 0.00 | (-0.14,0.13) | 0.07 | 0.07 | 0.062 | 0.00 | (-0.60,0.59) | 0.29 | 0.30 | 0.055 | 0.00 | 0.00 | 8.00 | 8 | 0.10 | 0.44 |
| | N1 | 0.01 | (-0.60,0.61) | 0.30 | 0.31 | 0.056 | 0.06 | (-2.46,2.58) | 1.27 | 1.29 | 0.055 | 1.00 | 0.00 | 8.01 | 7.96 | 0.17 | 0.36 |
| 3 | N2 | 0.01 | (-0.41,0.43) | 0.21 | 0.21 | 0.054 | 0.00 | (-1.73,1.74) | 0.91 | 0.88 | 0.037 | 1.00 | -0.01 | 8.01 | 8.01 | 0.18 | 0.37 |
| | N3 | 0.00 | (-0.14,0.13) | 0.07 | 0.07 | 0.058 | 0.01 | (-0.56,0.58) | 0.29 | 0.29 | 0.056 | 1.00 | 0.00 | 8.00 | 7.99 | 0.18 | 0.37 |
| | N1 | -0.02 | (-0.60,0.56) | 0.30 | 0.3 | 0.042 | 0.00 | (-2.55,2.54) | 1.27 | 1.30 | 0.051 | -0.01 | 0.02 | 8.00 | 8.01 | 0.18 | 0.37 |
| 1 | N2 | 0.02 | (-0.38,0.41) | 0.21 | 0.20 | 0.048 | 0.09 | (-1.7,1.87) | 0.90 | 0.91 | 0.057 | 0.01 | -0.01 | 8.01 | 7.93 | 0.18 | 0.37 |
| | N3 | 0.00 | (-0.13,0.13) | 0.07 | 0.07 | 0.054 | -0.01 | (-0.57,0.55) | 0.29 | 0.29 | 0.055 | 0.00 | 0.00 | 8.00 | 8.01 | 0.18 | 0.37 |

**Table A.3:** Bias of the treatment effect and variances after dropout for data simulated ($S = 1000$) under an outcome-dependent MNAR dropout mechanism according to DAG C. Data are simulated $S = 1000$ with intervention group mean and variance $\mu_1 = 1$ and $\sigma_1^2 = 8$), comparator group mean and variance $\mu_0 = 0$ and $\sigma_0^2 = 8$), 27% overall dropout, for sample sizes of $N1 = 500$, $N2 = 1000$ and $N3 = 10000$. We distinguish between data with normally distributed errors for $Y$ and $U$, and log-normally distributed errors. **1)** Strong logit dropout mechanism in the presence of a treatment effect ($\beta = 1$). **2)** Strong logit dropout mechanism under the null ($\beta = 0$). **3)** Weak logit dropout mechanism in the presence of a treatment effect ($\beta = 1$). **4)** Weak logit dropout mechanism under the null ($\beta = 0$). Shown are the bias and variance difference (VD), with 95% confidence confidence interval (CI), standard error (SE), Monte Carlo SE (MCSE), and proportion of 95% CIs excluding the null ($p_{95\%\text{CI}}$). Also included are the group means after dropout ( $\widehat{\mu}_1^*$, $\widehat{\mu}_0^*$), the group variances after dropout ($\widehat{\sigma}_0^{2*}$, $\widehat{\sigma}_1^{2*}$), and the dropout proportions per treatment group ($p_0$,$p_1$).

| | Size | Bias | 95% CI | SE | MCSE | $p_{95\%\text{CI}}$ | VD | 95% CI | SE | MCSE | $p_{95\%\text{CI}}$ | $\widehat{\mu}_1^*$ | $\widehat{\mu}_0^*$ | $\widehat{s}_1^{2*}$ | $\widehat{s}_0^{2*}$ | $p_1$ | $p_0$ |
|---|---|---|---|---|---|---|---|---|---|---|---|---|---|---|---|---|---|
| | | | | | | | Normally distributed errors | | | | | | | | | | |
| | N1 | -0.44 | (-0.85,-0.02) | 0.22 | 0.21 | 0.507 | 0.66 | (-0.70,2.02) | 0.70 | 0.69 | 0.156 | 2.02 | 1.46 | 4.80 | 4.14 | 0.22 | 0.33 |
| 1 | N2 | -0.44 | (-0.75,-0.12) | 0.16 | 0.16 | 0.785 | 0.68 | (-0.26,1.62) | 0.49 | 0.48 | 0.266 | 2.03 | 1.47 | 4.83 | 4.14 | 0.22 | 0.33 |
| | N3 | -0.44 | (-0.54,-0.34) | 0.05 | 0.05 | 1.00 | 0.68 | (0.39,0.97) | 0.16 | 0.15 | 0.993 | 2.02 | 1.46 | 4.82 | 4.14 | 0.22 | 0.33 |
| | N1 | 0.00 | (-0.49,0.48) | 0.24 | 0.25 | 0.05 | 0.02 | (-1.96,2.01) | 0.96 | 1.01 | 0.051 | 1.15 | 1.16 | 5.38 | 5.36 | 0.27 | 0.27 |
| 3 | N2 | 0.01 | (-0.32,0.34) | 0.17 | 0.17 | 0.042 | 0.04 | (-1.24,1.32) | 0.68 | 0.65 | 0.041 | 1.16 | 1.15 | 5.39 | 5.35 | 0.27 | 0.27 |
| | N3 | 0.00 | (-0.11,0.11) | 0.05 | 0.05 | 0.052 | 0.00 | (-0.41,0.41) | 0.22 | 0.21 | 0.037 | 1.15 | 1.16 | 5.38 | 5.38 | 0.27 | 0.27 |
| | N1 | -0.35 | (-0.82,0.12) | 0.24 | 0.24 | 0.316 | 0.48 | (-1.08,2.04) | 0.79 | 0.80 | 0.085 | 1.90 | 1.25 | 5.46 | 4.98 | 0.22 | 0.32 |
| 3 | N2 | -0.36 | (-0.70,-0.02) | 0.17 | 0.17 | 0.56 | 0.51 | (-0.59,1.62) | 0.56 | 0.56 | 0.157 | 1.89 | 1.25 | 5.48 | 4.96 | 0.22 | 0.32 |
| | N3 | -0.35 | (-0.45,-0.24) | 0.05 | 0.05 | 1.00 | 0.50 | (0.14,0.85) | 0.18 | 0.18 | 0.794 | 1.90 | 1.25 | 5.48 | 4.98 | 0.22 | 0.32 |
| | N1 | 0.00 | (-0.46,0.46) | 0.24 | 0.24 | 0.048 | 0.02 | (-1.53,1.57) | 0.78 | 0.79 | 0.051 | 1.09 | 1.09 | 5.18 | 5.16 | 0.27 | 0.27 |
| 4 | N2 | 0.00 | (-0.35,0.34) | 0.17 | 0.17 | 0.053 | 0.01 | (-1.06,1.07) | 0.56 | 0.55 | 0.038 | 1.10 | 1.1 | 5.17 | 5.16 | 0.27 | 0.27 |
| | N3 | 0.00 | (-0.10,0.10) | 0.05 | 0.05 | 0.045 | 0.00 | (-0.34,0.34) | 0.18 | 0.17 | 0.049 | 1.10 | 1.1 | 5.17 | 5.17 | 0.27 | 0.27 |
| | | | | | | | Log-normally distributed errors | | | | | | | | | | |
| | N1 | -0.48 | (-0.95,-0.01) | 0.25 | 0.24 | 0.483 | 0.66 | (-1.14,2.47) | 0.97 | 0.92 | 0.102 | 1.93 | 1.41 | 5.71 | 5.05 | 0.21 | 0.34 |
| 1 | N2 | -0.48 | (-0.82,-0.14) | 0.17 | 0.17 | 0.79 | 0.65 | (-0.72,2.01) | 0.69 | 0.70 | 0.153 | 1.93 | 1.41 | 5.72 | 5.07 | 0.21 | 0.34 |
| | N3 | -0.49 | (-0.59,-0.38) | 0.05 | 0.05 | 1.00 | 0.64 | (0.2,1.07) | 0.22 | 0.22 | 0.817 | 1.92 | 1.41 | 5.7 | 5.07 | 0.21 | 0.33 |
| | N1 | 0.00 | (-0.48,0.47) | 0.24 | 0.24 | 0.046 | -0.03 | (-1.98,1.92) | 0.95 | 0.99 | 0.064 | 1.15 | 1.15 | 5.35 | 5.37 | 0.27 | 0.27 |
| 2 | N2 | 0.01 | (-0.33,0.35) | 0.17 | 0.17 | 0.061 | -0.01 | (-1.43,1.42) | 0.68 | 0.73 | 0.07 | 1.16 | 1.15 | 5.37 | 5.37 | 0.27 | 0.27 |
| | N3 | 0.00 | (-0.10,0.10) | 0.05 | 0.05 | 0.041 | 0.00 | (-0.42,0.42) | 0.22 | 0.21 | 0.047 | 1.16 | 1.16 | 5.38 | 5.38 | 0.27 | 0.27 |
| | N1 | -0.36 | (-0.88,0.16) | 0.26 | 0.27 | 0.302 | 0.39 | (-1.74,2.53) | 1.04 | 1.09 | 0.07 | 1.84 | 1.21 | 6.32 | 5.93 | 0.22 | 0.33 |
| 3 | N2 | -0.38 | (-0.74,-0.01) | 0.18 | 0.19 | 0.532 | 0.43 | (-1.03,1.89) | 0.74 | 0.74 | 0.09 | 1.84 | 1.22 | 6.34 | 5.91 | 0.22 | 0.33 |
| | N3 | -0.38 | (-0.50,-0.26) | 0.06 | 0.06 | 1.00 | 0.40 | (-0.05,0.86) | 0.24 | 0.23 | 0.397 | 1.84 | 1.22 | 6.3 | 5.9 | 0.22 | 0.33 |
| | N1 | 0.01 | (-0.51,0.52) | 0.26 | 0.26 | 0.057 | 0.02 | (-2.09,2.12) | 1.04 | 1.07 | 0.053 | 1.02 | 1.01 | 6.09 | 6.08 | 0.27 | 0.27 |
| 4 | N2 | 0.00 | (-0.36,0.35) | 0.18 | 0.18 | 0.054 | -0.06 | (-1.54,1.42) | 0.74 | 0.75 | 0.053 | 1.02 | 1.02 | 6.07 | 6.13 | 0.27 | 0.27 |
| | N3 | 0.00 | (-0.12,0.12) | 0.06 | 0.06 | 0.065 | 0.00 | (-0.46,0.47) | 0.24 | 0.24 | 0.048 | 1.02 | 1.02 | 6.1 | 6.09 | 0.27 | 0.27 |

**Table A.4:** Bias of the treatment effect and variances after dropout for data simulated ($S = 1000$) under a non-outcome-dependent MNAR dropout mechanism according to DAG D (dropout dependent on treatment, $X$, and unmeasured variable, $U$, interacting on the log probability scale). Data are simulated $S = 1000$ with intervention group mean and variance $\mu_1 = 1$ and $\sigma_1^2 = 8$), comparator group mean and variance $\mu_0 = 0$ and $\sigma_0^2 = 8$), 27% overall dropout, for sample sizes of $N1 = 500$, $N2 = 1000$ and $N3 = 10000$. We distinguish between data with normally distributed errors for $Y$ and $U$, and log-normally distributed errors. **1)** Strong logit dropout mechanism in the presence of a treatment effect ($\beta = 1$). **2)** Strong logit dropout mechanism under the null ($\beta = 0$). **3)** Weak logit dropout mechanism in the presence of a treatment effect ($\beta = 1$). **4)** Weak logit dropout mechanism under the null ($\beta = 0$). Shown are the bias and variance difference (VD), with 95% confidence confidence interval (CI), standard error (SE), Monte Carlo SE (MCSE), and proportion of 95% CIs excluding the null ($p_{95\%\text{CI}}$). Also included are the group means after dropout ( $\widehat{\mu}_1^*$, $\widehat{\mu}_0^*$), the group variances after dropout ($\widehat{\sigma}_0^{2*}$, $\widehat{\sigma}_1^{2*}$), and the dropout proportions per treatment group ($p_0$,$p_1$).

| | Size | Bias | 95% CI | SE | MCSE | $p_{95\%\text{CI}}$ | VD | 95% CI | SE | MCSE | $p_{95\%\text{CI}}$ | $\widehat{\mu}_1^*$ | $\widehat{\mu}_0^*$ | $\widehat{s}_1^{2*}$ | $\widehat{s}_0^{2*}$ | $p_1$ | $p_0$ |
|---|---|---|---|---|---|---|---|---|---|---|---|---|---|---|---|---|---|
| | | | | | | | | Normally distributed errors | | | | | | | | | |
| | N1 | -0.39 | (-0.90,0.12) | 0.27 | 0.26 | 0.309 | 0.41 | (-1.40,2.22) | 0.95 | 0.92 | 0.068 | 1.64 | 1.03 | 6.58 | 6.17 | 0.20 | 0.35 |
| 1 | N2 | -0.41 | (-0.77,-0.04) | 0.19 | 0.19 | 0.57 | 0.43 | (-0.94,1.80) | 0.68 | 0.70 | 0.105 | 1.64 | 1.04 | 6.61 | 6.19 | 0.20 | 0.35 |
| | N3 | -0.4 | (-0.52,-0.29) | 0.06 | 0.06 | 1.00 | 0.44 | (0.02,0.86) | 0.22 | 0.21 | 0.521 | 1.64 | 1.04 | 6.61 | 6.18 | 0.20 | 0.35 |
| | N1 | -0.46 | (-0.98,0.07) | 0.28 | 0.27 | 0.38 | 0.31 | (-1.98,2.60) | 1.14 | 1.17 | 0.056 | 0.53 | 0.98 | 7.2 | 6.9 | 0.19 | 0.35 |
| 2 | N2 | -0.45 | (-0.84,-0.05) | 0.20 | 0.2 | 0.62 | 0.36 | (-1.26,1.97) | 0.82 | 0.82 | 0.075 | 0.53 | 0.98 | 7.22 | 6.87 | 0.18 | 0.35 |
| | N3 | -0.45 | (-0.57,-0.33) | 0.06 | 0.06 | 1.00 | 0.35 | (-0.16,0.85) | 0.26 | 0.26 | 0.268 | 0.53 | 0.98 | 7.21 | 6.86 | 0.19 | 0.35 |
| | N1 | -0.28 | (-0.81,0.24) | 0.28 | 0.27 | 0.163 | 0.25 | (-1.84,2.35) | 1.02 | 1.07 | 0.06 | 1.54 | 0.82 | 7.02 | 6.78 | 0.21 | 0.33 |
| 3 | N2 | -0.27 | (-0.65,0.11) | 0.20 | 0.19 | 0.261 | 0.28 | (-1.11,1.67) | 0.72 | 0.71 | 0.065 | 1.55 | 0.82 | 7.02 | 6.75 | 0.21 | 0.33 |
| | N3 | -0.28 | (-0.39,-0.16) | 0.06 | 0.06 | 0.997 | 0.25 | (-0.20,0.71) | 0.23 | 0.23 | 0.202 | 1.54 | 0.82 | 7.02 | 6.77 | 0.21 | 0.33 |
| | N1 | -0.28 | (-0.82,0.27) | 0.28 | 0.28 | 0.161 | 0.34 | (-1.63,2.32) | 1.02 | 1.01 | 0.058 | 0.54 | 0.82 | 7.04 | 6.7 | 0.21 | 0.33 |
| 4 | N2 | -0.28 | (-0.65,0.10) | 0.20 | 0.19 | 0.289 | 0.28 | (-1.11,1.68) | 0.72 | 0.71 | 0.06 | 0.54 | 0.81 | 7.03 | 6.75 | 0.21 | 0.33 |
| | N3 | -0.28 | (-0.40,-0.15) | 0.06 | 0.06 | 0.993 | 0.26 | (-0.18,0.71) | 0.23 | 0.23 | 0.197 | 0.54 | 0.82 | 7.02 | 6.76 | 0.21 | 0.33 |
| | | | | | | | | Log-normally distributed errors | | | | | | | | | |
| | N1 | -0.45 | (-1.00,0.10) | 0.28 | 0.28 | 0.354 | 0.33 | (-1.98,2.64) | 1.15 | 1.18 | 0.054 | 1.53 | 0.98 | 7.18 | 6.86 | 0.19 | 0.35 |
| 1 | N2 | -0.45 | (-0.84,-0.07) | 0.20 | 0.2 | 0.603 | 0.40 | (-1.21,2) | 0.82 | 0.82 | 0.076 | 1.53 | 0.98 | 7.25 | 6.86 | 0.19 | 0.35 |
| | N3 | -0.45 | (-0.57,-0.33) | 0.06 | 0.06 | 1.00 | 0.36 | (-0.16,0.87) | 0.26 | 0.26 | 0.276 | 1.53 | 0.98 | 7.21 | 6.86 | 0.19 | 0.35 |
| | N1 | -0.46 | (-1.00,0.08) | 0.28 | 0.28 | 0.363 | 0.34 | (-1.87,2.56) | 1.15 | 1.13 | 0.058 | 0.53 | 0.98 | 7.22 | 6.88 | 0.19 | 0.35 |
| 2 | N2 | -0.45 | (-0.82,-0.08) | 0.20 | 0.19 | 0.637 | 0.35 | (-1.23,1.93) | 0.81 | 0.81 | 0.066 | 0.53 | 0.98 | 7.22 | 6.88 | 0.19 | 0.35 |
| | N3 | -0.46 | (-0.58,-0.33) | 0.06 | 0.06 | 1.00 | 0.33 | (-0.18,0.84) | 0.26 | 0.26 | 0.247 | 0.53 | 0.98 | 7.21 | 6.88 | 0.19 | 0.35 |
| | N1 | -0.31 | (-0.87,0.26) | 0.29 | 0.29 | 0.195 | 0.12 | (-2.26,2.49) | 1.20 | 1.21 | 0.043 | 1.46 | 0.77 | 7.53 | 7.42 | 0.20 | 0.33 |
| 3 | N2 | -0.3 | (-0.69,0.09) | 0.20 | 0.2 | 0.321 | 0.17 | (-1.52,1.86) | 0.85 | 0.86 | 0.056 | 1.47 | 0.77 | 7.57 | 7.41 | 0.20 | 0.34 |
| | N3 | -0.3 | (-0.43,-0.18) | 0.06 | 0.07 | 0.999 | 0.12 | (-0.41,0.66) | 0.27 | 0.27 | 0.071 | 1.46 | 0.77 | 7.56 | 7.43 | 0.20 | 0.34 |
| | N1 | -0.3 | (-0.87,0.27) | 0.29 | 0.29 | 0.18 | 0.13 | (-2.32,2.58) | 1.20 | 1.25 | 0.059 | 0.46 | 0.76 | 7.58 | 7.46 | 0.20 | 0.33 |
| 3 | N2 | -0.3 | (-0.72,0.11) | 0.20 | 0.21 | 0.333 | 0.16 | (-1.52,1.85) | 0.85 | 0.86 | 0.053 | 0.46 | 0.76 | 7.57 | 7.41 | 0.20 | 0.33 |
| | N3 | -0.3 | (-0.43,-0.18) | 0.06 | 0.06 | 0.997 | 0.13 | (-0.42,0.67) | 0.27 | 0.28 | 0.08 | 0.46 | 0.77 | 7.56 | 7.43 | 0.20 | 0.33 |

**Table A.5:** Bias of the treatment effect and variances after dropout for data simulated ($S = 1000$) under an MNAR dropout mechanism according to DAG E, where dropout depends on some unmeasured variable, $U$. Data are simulated $S = 1000$ with intervention group mean and variance $\mu_1 = 1$ and $\sigma_1^2 = 8$), comparator group mean and variance $\mu_0 = 0$ and $\sigma_0^2 = 8$), 27% overall dropout, for sample sizes of $N1 = 500$, $N2 = 1000$ and $N3 = 10000$. We distinguish between data with normally distributed errors for $Y$ and $U$, and log-normally distributed errors. **1)** Strong logit dropout mechanism in the presence of a treatment effect ($\beta = 1$). **2)** Strong logit dropout mechanism under the null ($\beta = 0$). **3)** Weak logit dropout mechanism in the presence of a treatment effect ($\beta = 1$). **4)** Weak logit dropout mechanism under the null ($\beta = 0$). Shown are the bias and variance difference (VD), with 95% confidence confidence interval (CI), standard error (SE), Monte Carlo SE (MCSE), and proportion of 95% CIs excluding the null ($p_{95\%\text{CI}}$). Also included are the group means after dropout ( $\widehat{\mu}_1^*$, $\widehat{\mu}_0^*$), the group variances after dropout ($\widehat{\sigma}_0^{2*}$, $\widehat{\sigma}_1^{2*}$), and the dropout proportions per treatment group ($p_0$,$p_1$).

| | Size | Bias | 95% CI | SE | MCSE | $p_{95\%\text{CI}}$ | VD | 95% CI | SE | MCSE | $p_{95\%\text{CI}}$ | $\widehat{\mu}_1^*$ | $\widehat{\mu}_0^*$ | $\widehat{s}_1^{2*}$ | $\widehat{s}_0^{2*}$ | $p_1$ | $p_0$ |
|---|---|---|---|---|---|---|---|---|---|---|---|---|---|---|---|---|---|
| | | | | | | | Normally distributed errors | | | | | | | | | | |
| | N1 | 0.01 | (-0.51,0.52) | 0.26 | 0.26 | 0.048 | -0.01 | (-1.89,1.87) | 0.94 | 0.96 | 0.05 | 1.82 | 0.82 | 6.38 | 6.39 | 0.27 | 0.27 |
| 1 | N2 | 0.00 | (-0.37,0.38) | 0.19 | 0.19 | 0.052 | 0.03 | (-1.33,1.39) | 0.67 | 0.70 | 0.062 | 1.83 | 0.82 | 6.4 | 6.37 | 0.27 | 0.27 |
| | N3 | 0.00 | (-0.11,0.12) | 0.06 | 0.06 | 0.049 | 0.01 | (-0.40,0.41) | 0.21 | 0.21 | 0.038 | 1.83 | 0.82 | 6.39 | 6.39 | 0.27 | 0.27 |
| | N1 | 0.00 | (-0.54,0.53) | 0.28 | 0.27 | 0.047 | 0.03 | (-2.17,2.23) | 1.13 | 1.12 | 0.057 | 0.75 | 0.76 | 6.99 | 6.96 | 0.27 | 0.27 |
| 2 | N2 | 0.01 | (-0.38,0.40) | 0.20 | 0.2 | 0.065 | 0.01 | (-1.67,1.68) | 0.81 | 0.85 | 0.058 | 0.77 | 0.76 | 7.02 | 7.01 | 0.27 | 0.27 |
| | N3 | 0.00 | (-0.12,0.13) | 0.06 | 0.06 | 0.048 | 0.01 | (-0.51,0.52) | 0.26 | 0.26 | 0.054 | 0.76 | 0.76 | 7.02 | 7.01 | 0.27 | 0.27 |
| | N1 | -0.01 | (-0.54,0.52) | 0.28 | 0.27 | 0.046 | 0.04 | (-2.04,2.11) | 1.01 | 1.06 | 0.063 | 1.69 | 0.70 | 6.89 | 6.86 | 0.27 | 0.27 |
| 3 | N2 | -0.01 | (-0.39,0.38) | 0.19 | 0.20 | 0.055 | -0.03 | (-1.44,1.39) | 0.72 | 0.72 | 0.042 | 1.69 | 0.70 | 6.87 | 6.9 | 0.27 | 0.27 |
| | N3 | 0.00 | (-0.12,0.12) | 0.06 | 0.06 | 0.052 | 0.00 | (-0.45,0.44) | 0.23 | 0.23 | 0.045 | 1.69 | 0.69 | 6.86 | 6.87 | 0.27 | 0.27 |
| | N1 | 0.00 | (-0.57,0.56) | 0.28 | 0.29 | 0.063 | 0.04 | (-1.95,2.03) | 1.01 | 1.02 | 0.053 | 0.69 | 0.69 | 6.88 | 6.84 | 0.27 | 0.27 |
| 4 | N2 | 0.00 | (-0.39,0.38) | 0.19 | 0.20 | 0.055 | 0.02 | (-1.45,1.48) | 0.72 | 0.75 | 0.056 | 0.69 | 0.69 | 6.87 | 6.86 | 0.27 | 0.27 |
| | N3 | 0.00 | (-0.12,0.12) | 0.06 | 0.06 | 0.04 | 0.02 | (-0.43,0.47) | 0.23 | 0.23 | 0.055 | 0.69 | 0.69 | 6.88 | 6.86 | 0.27 | 0.27 |
| | | | | | | | Log-normally distributed errors | | | | | | | | | | |
| | N1 | -0.01 | (-0.54,0.52) | 0.28 | 0.27 | 0.034 | -0.04 | (-2.25,2.18) | 1.13 | 1.13 | 0.045 | 1.75 | 0.76 | 6.99 | 7.02 | 0.27 | 0.27 |
| 1 | N2 | 0.00 | (-0.38,0.37) | 0.20 | 0.19 | 0.047 | -0.02 | (-1.60,1.56) | 0.81 | 0.81 | 0.051 | 1.76 | 0.76 | 7.01 | 7.03 | 0.27 | 0.27 |
| | N3 | 0.00 | (-0.12,0.12) | 0.06 | 0.06 | 0.046 | 0.00 | (-0.51,0.51) | 0.26 | 0.26 | 0.05 | 1.76 | 0.76 | 7.02 | 7.02 | 0.27 | 0.27 |
| | N1 | 0.00 | (-0.56,0.55) | 0.28 | 0.28 | 0.055 | 0.02 | (-2.18,2.21) | 1.13 | 1.12 | 0.053 | 0.76 | 0.76 | 7.04 | 7.03 | 0.27 | 0.27 |
| 2 | N2 | 0.01 | (-0.39,0.40) | 0.20 | 0.2 | 0.049 | 0.05 | (-1.62,1.71) | 0.81 | 0.85 | 0.062 | 0.76 | 0.76 | 7.05 | 7.01 | 0.27 | 0.27 |
| | N3 | 0.00 | (-0.12,0.13) | 0.06 | 0.06 | 0.044 | -0.01 | (-0.51,0.49) | 0.26 | 0.26 | 0.048 | 0.76 | 0.76 | 7.01 | 7.02 | 0.27 | 0.27 |
| | N1 | -0.01 | (-0.57,0.56) | 0.29 | 0.29 | 0.051 | 0.02 | (-2.23,2.26) | 1.19 | 1.14 | 0.044 | 1.61 | 0.62 | 7.46 | 7.45 | 0.27 | 0.27 |
| 3 | N2 | 0.00 | (-0.40,0.40) | 0.20 | 0.2 | 0.056 | 0.02 | (-1.65,1.69) | 0.85 | 0.85 | 0.046 | 1.62 | 0.62 | 7.49 | 7.47 | 0.27 | 0.27 |
| | N3 | 0.00 | (-0.13,0.13) | 0.06 | 0.07 | 0.061 | -0.02 | (-0.55,0.52) | 0.27 | 0.27 | 0.049 | 1.62 | 0.62 | 7.48 | 7.49 | 0.27 | 0.27 |
| | N1 | -0.01 | (-0.57,0.55) | 0.29 | 0.29 | 0.053 | -0.03 | (-2.35,2.30) | 1.20 | 1.18 | 0.041 | 0.62 | 0.63 | 7.48 | 7.5 | 0.27 | 0.27 |
| 4 | N2 | 0.01 | (-0.39,0.41) | 0.20 | 0.2 | 0.048 | -0.03 | (-1.70,1.64) | 0.85 | 0.85 | 0.056 | 0.62 | 0.61 | 7.46 | 7.5 | 0.27 | 0.27 |
| | N3 | 0.00 | (-0.12,0.13) | 0.06 | 0.06 | 0.043 | 0.00 | (-0.52,0.52) | 0.27 | 0.27 | 0.055 | 0.62 | 0.62 | 7.48 | 7.48 | 0.27 | 0.27 |

### Appendix B

This appendix gives a detailed description and the results of the simulation study of Section 6 of the main text in full. Section B.1 describes the simulation approach in detail. Section B.2 gives results for a simulation with balanced randomization of patients to treatment groups, with Table B.1 an extended companion table to Table 1 of the main text. Section B.3 gives results for a simulation with unbalanced randomization, with Table B.2 designed analogously to Table 1 of the main text, and Table B.3 a companion table to Table B.2.

#### B.1 Simulation approach

Here, we describe the simulation approach in detail using the ADEMP framework, and define the aims, data-generating mechanisms, estimands, methods, and performance measures.

**Aim**: Illustrating that, for longitudinal data, 1) a variance difference in outcomes at follow-up may be the result of outcome-dependent dropout, non-outcome dependent dropout or treatment heterogeneity (resulting from effect modification); 2) a variance difference in outcomes at baseline only results from outcome-dependent dropout; 3) in the presence of effect modification, conditioning on the effect modifier (EM) and the treatment, $X$, and EM interaction term, results in a zero outcome variance difference at follow-up.

**General setup:** Longitudinal data with correlated outcomes at baseline and follow-up, drawn from a multivariate normal distribution, simulated at a sample size of $N = 1000$, with balanced and unbalanced randomization to intervention and comparator group and a positive treatment effect, for four different dropout scenarios, simulated using a logit mechanism.

<u>Data generating mechanism:</u>

**Variables**

1. Binary treatment variable, $X \sim B(1000, 0.5)$, with half of 1000 patients randomized to the intervention group (X=1) and half to the comparator group (X=0), some normally distributed variable $U \sim N(0, 1)$, which affects the outcome at final follow-up, $Y_f$, and outcomes at baseline ($Y_b$) and final follow-up drawn from a multivariate normal distribution, and a positive treatment effect of $\beta = 1$, with

$$Y_f = X + U + \varepsilon_f$$

and

$$\begin{pmatrix} Y_b \\ \varepsilon_f \end{pmatrix} \sim N\left(\begin{pmatrix} 0 \\ 0 \end{pmatrix}, \begin{pmatrix} 4 & 2.9 \\ 2.9 & 4 \end{pmatrix}\right),$$

so that $\text{var}(Y_b) = 4$ and $\text{var}(Y_f) = \text{var}(U + \varepsilon_f) = 5$, with a correlation coefficient $\text{cor}(Y_b, Y_f) = 0.65$. Results are reported in the main text, Table 1 and in Supplementary Table B.1).

2. Identical setup, but with treatment variable $X \sim B(1, 0.35)$, so that approximately 35% of 1000 patients are randomized to the intervention group. Results are reported in Supplementary Tables B.2 and B.3).

Each of the two scenarios is considered without effect modification, and with effect modification, where $Y_b$ acts as a positive effect modifier in the intervention group:

$$Y_f = X + U + 0.5Y_bX + \varepsilon_f.$$

**Dropout mechanism:** Logit selection mechanism with intercepts selected such that overall dropout proportion is approximately 0.27, with four different dropout scenarios considered (shown as directed acyclic graphs (DAGs) A to D in Figure 1 of the main text). For each scenario, the probability of selection is defined conditional on the model variables: $\mathsf{P}(R = 1|Y, X, U)$, with $R$ the response indicator.

A *No dropout*
With and without effect modification

- $\mathsf{P}(R = 1|Y, X, U) = 1$

B *$X$-dependent dropout*
With and without effect modification

- $\mathsf{P}(R = 1|Y, X, U) = \exp(0.2 + 2X)/(1 + \exp(0.2 + 2X)$

C *$Y$-dependent dropout*
With effect modification

- $\mathsf{P}(R = 1|Y, X, U) = \exp(5.2 - 2.5Y)/(1 + \exp(5.2 - 2.5Y)$

Without effect modification

- $\mathsf{P}(R = 1|Y, X, U) = \exp(4.8 - 2.5Y)/(1 + \exp(4.8 - 2.5Y)$

D *Non-$Y$-dependent MNAR dropout ($X$- and $U$-dependent*
With and without effect modification

- $\mathsf{P}(R = 1|Y, X, U) = \exp(0.7 + 2X + 2U)/(1 + \exp(0.7 + 2X + 2U)$

**Simulation size:** For each scenario, 1000 datasets were simulated. Simulation quality was verified by checking of mean SEs of the CCA and variance difference estimates were comparable to the Monte Carlo SEs (see 'Performance measures')

**Estimands**
*Primary:* Reported in main text, Figure 3, and in appendix, Supplementary Figures 1 and 2.

- Bias of the complete case analysis (CCA) treatment effect estimate, regressing outcome, $Y_f$, on treatment, $X$, with 95% CI

- Bias of the adjusted CCA treatment effect estimate, regressing outcome, $Y_f$, on treatment, $X$, and the outcome at baseline, $Y_b$, with 95% CI
- Unadjusted outcome variance difference at follow-up, with 95% CI
- Outcome variance difference at follow-up adjusted for outcome at baseline, $Y_b$, with 95% CI
- Outcome variance difference at follow-up adjusted for outcome at baseline, $Y_b$ and the interaction term between treatment, $X$ and $Y_b$, with 95% CI
- Unadjusted outcome variance difference at baseline, with 95% CI

*Secondary:* Reported in appendix, Tables A1 to A5, for dropout scenarios A to E.

- 95% coverage* of all six measures listed above
- Standard error (SE) of all six measures listed above
- Monte Carlo SE (MCSE) of all six measures listed above

(* The 95% coverage refers to the proportion of times the estimate was excluded from the 95% CI. If the true estimate is 0, this should come to 0.05)

**Methods**

- CCA estimator
- Studentized Breusch-Pagan test (for estimating the variance difference across groups after dropout)
- 95% CIs were calculated using the Monte Carlo Standard error (MCSE)

**Performance measures**: Simulation quality was checked by seeing if the estimate SEs were comparable to the MCSEs

#### B.2 Simulation results: balanced randomization

Table B.1 is a companion table to Table 1 in the main text, and additionally reports, for each estimate, the standard error (SE), the Monte Carlo SE (MCSE), and the proportion of 95% CIs excluding the null ($p_{95\%CI}$). We observe that the SEs and MCSEs are comparable, suggesting that a simulation size of 1000 is sufficient.

**Table B.1:** Companion table to Table B.2. Bias of the complete case analysis (CCA) treatment effect estimate and variance difference (VD) in the observed sample with 95% confidence intervals (CIs), standard error (SE), Monte Carlo SE (MCSE) and proportion of 95% CIs excluding the null ($p_{95\%CI}$), for longitudinal data, with baseline ($Y_b$) and follow-up ($Y_f$) measurements, simulated under different dropout mechanisms, without and with effect modification (EM). Of 1000 patients, 50% were randomized to intervention and 50% to control. Estimates were obtained without and with conditioning on the baseline outcome, with VDs calculated at baseline ($\mathrm{VD}_b$) and at final follow-up ($\mathrm{VD}_f$), for the set of patients still observed at final follow-up. VD at final follow-up was also calculated conditional on $Y_b$ and the interaction between $Y_b$ and treatment ($\mathrm{VD}_{f(bI)}$). A) No dropout in both groups; B) MAR dropout dependent on treatment; C) MNAR dropout dependent on outcome, $Y_f$; D) MNAR dropout dependent on treatment and an unmeasured covariate, $U$, interacting on the probability scale.

| | *Unadjusted for outcome at baseline* | | | | | | | | | | | | *Adjusted for outcome at baseline* | | | | | | | | | | | |
|---|---|---|---|---|---|---|---|---|---|---|---|---|---|---|---|---|---|---|---|---|---|---|---|---|
| | Bias (95% CI) | SE | MCSE | $p_{95\%CI}$ | $\mathrm{VD}_b$ (95% CI) | SE | MCSE | $p_{95\%CI}$ | $\mathrm{VD}_f$ (95% CI) | SE | MCSE | $p_{95\%CI}$ | Bias (95% CI) | SE | MCSE | $p_{95\%CI}$ | $\mathrm{VD}_{f(b)}$ (95% CI) | SE | MCSE | $p_{95\%CI}$ | $\mathrm{VD}_{f(bI)}$ (95% CI) | SE | MCSE | $p_{95\%CI}$ |
| *No dropout* | | | | | | | | | | | | | *No dropout* | | | | | | | | | | | |
| A | 0.00 (-0.29,0.29) | 0.15 | 0.15 | 0.053 | 0.01 (-0.70,0.72) | 0.37 | 0.36 | 0.041 | 0.01 (-0.90,0.93) | 0.47 | 0.47 | 0.05 | 0.00 (-0.22,0.22) | 0.11 | 0.11 | 0.048 | 0.01 (-0.52,0.54) | 0.27 | 0.27 | 0.048 | 0.01 (-0.53,0.54) | 0.27 | 0.27 | 0.05 |
| A(EM) | 0.00 (-0.37,0.37) | 0.17 | 0.19 | 0.079 | -0.01 (-0.72,0.70) | 0.37 | 0.36 | 0.046 | 3.90 (2.5,5.30) | 0.62 | 0.71 | 1.00 | 0.00 (-0.24,0.24) | 0.12 | 0.12 | 0.063 | 0.29 (-0.25,0.84) | 0.29 | 0.28 | 0.155 | -0.01 (-0.54,0.52) | 0.27 | 0.27 | 0.051 |
| *MAR dropout* | | | | | | | | | | | | | *MAR dropout* | | | | | | | | | | | |
| B | 0.00 (-0.32,0.33) | 0.17 | 0.17 | 0.05 | 0.00 (-0.82,0.81) | 0.42 | 0.41 | 0.047 | 0.00 (-1.02,1.03) | 0.53 | 0.52 | 0.048 | 0.00 (-0.25,0.24) | 0.13 | 0.12 | 0.043 | 0.01 (-0.59,0.61) | 0.30 | 0.31 | 0.053 | 0.01 (-0.59,0.61) | 0.30 | 0.31 | 0.053 |
| B(EM) | 0.00 (-0.40,0.40) | 0.19 | 0.20 | 0.067 | 0.00 (-0.80,0.80) | 0.42 | 0.41 | 0.038 | 3.87 (2.35,5.38) | 0.73 | 0.77 | 1.00 | 0.00 (-0.26,0.26) | 0.13 | 0.13 | 0.041 | 0.11 (-0.49,0.70) | 0.33 | 0.31 | 0.044 | -0.01 (-0.60,0.59) | 0.30 | 0.3 | 0.047 |
| *Outcome-dependent MNAR dropout* | | | | | | | | | | | | | *Outcome-dependent MNAR dropout* | | | | | | | | | | | |
| C | -0.45 (-0.71,-0.18) | 0.14 | 0.14 | 0.91 | -0.18 (-0.93,0.56) | 0.37 | 0.38 | 0.085 | -0.54 (-1.15,0.08) | 0.33 | 0.31 | 0.37 | -0.32 (-0.54,-0.09) | 0.11 | 0.11 | 0.784 | -0.27 (-0.72,0.17) | 0.23 | 0.23 | 0.215 | -0.28 (-0.73,0.16) | 0.23 | 0.23 | 0.234 |
| C(EM) | -1.03 (-1.34,-0.72) | 0.15 | 0.16 | 1.00 | -0.81 (-0.93,0.56) | 0.36 | 0.32 | 0.625 | 1.04 (0.1,1.99) | 0.41 | 0.48 | 0.694 | -0.7 (-0.93,-0.46) | 0.12 | 0.12 | 1.00 | 0.15 (-0.36,0.66) | 0.26 | 0.26 | 0.099 | -0.02 (-0.50,0.46) | 0.25 | 0.24 | 0.051 |
| *Non-outcome-dependent MNAR dropout* | | | | | | | | | | | | | *Non-outcome-dependent MNAR dropout* | | | | | | | | | | | |
| D | -0.25 (-0.59,0.08) | 0.16 | 0.17 | 0.337 | 0.01 (-0.83,0.85) | 0.43 | 0.43 | 0.058 | 0.13 (-0.87,1.13) | 0.50 | 0.51 | 0.061 | -0.25 (-0.50,0.00) | 0.12 | 0.13 | 0.525 | 0.13 (-0.44,0.69) | 0.28 | 0.29 | 0.086 | 0.12 (-0.44,0.69) | 0.28 | 0.29 | 0.086 |
| D(EM) | -0.26 (-0.65,0.13) | 0.19 | 0.20 | 0.291 | 0.01 (-0.84,0.86) | 0.42 | 0.43 | 0.06 | 4.00 (2.43,5.58) | 0.70 | 0.80 | 1.00 | -0.26 (-0.52,-0.01) | 0.13 | 0.13 | 0.541 | 0.28 (-0.28,0.84) | 0.30 | 0.29 | 0.141 | 0.12 (-0.43,0.67) | 0.28 | 0.28 | 0.071 |

#### B.3 Simulation results: unbalanced randomization

In Section 6 of the main text, we considered the effect of treatment effect heterogeneity, resulting from effect modification (EM), on the variance difference at baseline and at final follow-up, for longitudinal data. We showed that outcome-dependent dropout results in a variance difference at baseline ($\text{VD}_b$) and at final follow-up ($\text{VD}_f$) both, while treatment effect heterogeneity results in a variance difference at final follow-up only. Adjusting for the outcome at baseline, $Y_b$, resulted in a decreased variance difference ($\text{VD}_{f(b)}$). We also showed that when patients are randomized to equally sized treatment groups, additionally adjusting for the interaction between treatment and $Y_b$ left the variance term difference estimate ($\text{VD}_{f(bI)}$) unchanged in the absence of dropout, but resulted in different variance differences (both smaller and larger) under differential dropout. Here we consider unequal treatment groups, with 35% of patients randomized to intervention and 65% randomized to treatment. The presence of treatment effect heterogeneity was assessed by testing if severity at baseline acts as an effect modifier. This was done by calculating the variance difference estimate adjusted for $Y_b$ and the interaction between $Y_b$ and treatment, $X$ ($\text{VD}_{f(bI)}$). In Table B.2 we observe that when treatment groups are unequal, conditioning on the $Y_b$ and treatment interaction term results in a decreased variance difference only when $Y_b$ acts as an effect modifier (scenarios 1B-4B). Table B.3 is a companion table to Table B.2, and additionally reports, for each estimate, the standard error (SE), the Monte Carlo SE (MCSE), and the proportion of 95% CIs excluding the null ($p_{95\%CI}$). We observe comparable SEs and MCSEs, suggesting that a simulation size of 1000 is sufficient.

**Table B.2:** Bias of the complete case analysis (CCA) treatment effect estimate and variance difference (VD) in the observed sample with 95% confidence intervals (CIs), for longitudinal data, with baseline ($Y_b$) and follow-up ($Y_f$) measurements, simulated under different dropout mechanisms, without and with effect modification (EM). Of 1000 patients, 35% were randomized to intervention and 65% to control. Estimates were obtained without and with conditioning on the baseline outcome, with VDs calculated at baseline ($\text{VD}_b$) and at final follow-up ($\text{VD}_f$), for the set of patients still observed at final follow-up. VD at final follow-up was also calculated conditional on $Y_b$ ($\text{VD}_{f(b)}$) and conditional on $Y_b$ and the interaction between $Y_b$ and treatment ($\text{VD}_{f(bI)}$). A) No dropout in both groups; B) MAR dropout dependent on treatment; C) MNAR dropout dependent on outcome, $Y_f$; D) MNAR dropout dependent on treatment and an unmeasured covariate, $U$, interacting on the probability scale.

| | *Unadjusted for outcome at baseline* | | | *Adjusted for outcome at baseline* | | |
|---|---|---|---|---|---|---|
| | Bias (95% CI) | $\text{VD}_b$ (95% CI) | $\text{VD}_f$ (95% CI) | Bias (95% CI) | $\text{VD}_{f(b)}$ (95% CI) | $\text{VD}_{f(bI)}$ (95% CI) |
| *No dropout* | | | | | | |
| A | 0.00 (-0.29,0.29) | 0.01 (-0.70,0.72) | 0.01 (-0.90,0.93) | 0.00 (-0.22,0.22) | 0.01 (-0.52,0.54) | 0.01 (-0.53,0.54) |
| A(EM) | 0.00 (-0.37,0.37) | -0.01 (-0.72,0.70) | 3.90 (2.50,5.30) | 0.00 (-0.24,0.24) | 0.29 (-0.25,0.84) | -0.01 (-0.54,0.52) |
| *MAR dropout* | | | | | | |
| B | 0.00 (-0.32,0.33) | 0.00 (-0.82,0.81) | 0.00 (-1.02,1.03) | 0.00 (-0.25,0.24) | 0.01 (-0.59,0.61) | 0.01 (-0.59,0.61) |
| B(EM) | 0.00 (-0.40,0.40) | 0.00 (-0.80,0.80) | 3.87 (2.35,5.38) | 0.00 (-0.26,0.26) | 0.11 (-0.49,0.70) | -0.01 (-0.60,0.59) |
| *Ooutcome-dependent MNAR dropout* | | | | | | |
| C | -0.45 (-0.71,-0.18) | -0.18 (-0.93,0.56) | -0.54 (-1.15,0.08) | -0.32 (-0.54,-0.09) | -0.27 (-0.72,0.17) | -0.28 (-0.73,0.16) |
| C(EM) | -1.03 (-1.34,-0.72) | -0.81 (-0.93,0.56) | 1.04 (0.10,1.99) | -0.70 (-0.93,-0.46) | 0.15 (-0.36,0.66) | -0.02 (-0.50,0.46) |
| *Non-outcome-dependent MNAR dropout* | | | | | | |
| D | -0.25 (-0.59,0.08) | 0.01 (-0.83,0.85) | 0.13 (-0.87,1.13) | -0.25 (-0.50,0.00) | 0.13 (-0.44,0.69) | 0.12 (-0.44,0.69) |
| D(EM) | -0.26 (-0.65,0.13) | 0.01 (-0.84,0.86) | 4.00 (2.43,5.58) | -0.26 (-0.52,-0.01) | 0.28 (-0.28,0.84) | 0.12 (-0.43,0.67) |

### Appendix C

This appendix gives a detailed description and the results of the simulation study of Section 7 of the main text in full. Section C.1 describes the simulation approach in detail. Section C.2 gives results for a simulation at sample sizes of $N = 1000$ and $N = 10000$, Table C.1 a companion table to Table 2 of the main text.

#### C.1 Simulation approach

Here, we describe the simulation approach in detail using the ADEMP framework, and define the aims, data-generating mechanisms, estimands, methods, and performance measures.

**Aim**: Illustrating that 1) when performing a multiple imputation (MI) analysis, the variance difference may be used to assess the added value of including variables in the imputation model; 2) if dropout is MAR conditional on the imputation model covariates, then the treatment effect, estimated with the analysis model is unbiased and the variance difference, conditional on the analysis model (close to) zero; 3) if dropout is MNAR conditional on the imputation model covariates, then the treatment effect, estimated with the analysis model is biased and the variance difference, conditional on the analysis model non-zero.

**General setup:** Data with normally distributed outcomes, simulated with a treatment effect of $\beta = 1$, at two different sample sizes, for two different dropout scenarios, simulated with a logit mechanism.

Data generating mechanism:

**Variables**: Binary treatment variable, $X \sim B(1, 0.5)$, some normally distributed variables $A \sim N(0, 2)$ and $B \sim N(0, 2)$, which affect $Y$, the outcome at final follow-up, so that $Y = \beta X + A + B + \varepsilon$, with a positive treatment effect of $\beta = 1$, and $\varepsilon \sim N(0, 4)$, so that $\text{var}(Y|X) = 8$. (Results reported in main text, Table 2, and in Appendix C.2, Table C.1)

**Sample size**: Total sample sizes of $N = 1000$ and $N = 10000$

**Dropout mechanism:** Logit selection mechanism with intercepts selected such that overall dropout proportion is approximately 0.27, with two different dropout scenarios considered (shown as directed acyclic graphs (DAGs) M1 and M2 in Figure 5 of the main text). For each scenario, the probability of selection is defined conditional on the model variables: $\mathsf{P}(R = 1|Y, X, A, B)$, with $R$ the response indicator.

M1 *MAR dropout dependent on $B$ and $X$*

- $\mathsf{P}(R = 1|Y, X, A, B) = \exp(1.1 + 2.1X)/(1 + \exp(1.1 + 2.2X)$

**Table B.3:** Companion table to Table B.2. Bias of the complete case analysis (CCA) treatment effect estimate and variance difference (VD) in the observed sample with 95% confidence intervals (CIs), standard error (SE), Monte Carlo SE (MCSE) and proportion of 95% CIs excluding the null ($p_{95\%CI}$), for longitudinal data, with baseline ($Y_b$) and follow-up ($Y_f$) measurements, simulated under different dropout mechanisms, without and with effect modification (EM). Of 1000 patients, 35% were randomized to intervention and 65% to control. Estimates were obtained without and with conditioning on the baseline outcome, with VDs calculated at baseline ($\text{VD}_b$) and at final follow-up ($\text{VD}_f$), for the set of patients still observed at final follow-up. VD at final follow-up was also calculated conditional on $Y_b$ and the interaction between $Y_b$ and treatment ($\text{VD}_{f(bI)}$). A) No dropout in both groups; B) MAR dropout dependent on treatment; C) MNAR dropout dependent on outcome, $Y_f$; D) MNAR dropout dependent on treatment and an unmeasured covariate, $U$, interacting on the probability scale.

| | *Unadjusted for outcome at baseline* | | | | | | | | | | | | *Adjusted for outcome at baseline* | | | | | | | | | | | |
|---|---|---|---|---|---|---|---|---|---|---|---|---|---|---|---|---|---|---|---|---|---|---|---|---|
| | Bias (95% CI) | SE | MCSE | $p_{95\%CI}$ | $\text{VD}_b$ (95% CI) | SE | MCSE | $p_{95\%CI}$ | $\text{VD}_f$ (95% CI) | SE | MCSE | $p_{95\%CI}$ | Bias (95% CI) | SE | MCSE | $p_{95\%CI}$ | $\text{VD}_{f(b)}$ (95% CI) | SE | MCSE | $p_{95\%CI}$ | $\text{VD}_{f(bI)}$ (95% CI) | SE | MCSE | $p_{95\%CI}$ |
| *No dropout* | | | | | | | | | | | | | *No dropout* | | | | | | | | | | | |
| A | 0.00 (-0.28,0.27) | 0.14 | 0.14 | 0.046 | 0.00 (-0.71,0.72) | 0.36 | 0.37 | 0.06 | 0.01 (-0.88,0.9) | 0.45 | 0.45 | 0.046 | 0.00 (-0.22,0.22) | 0.11 | 0.11 | 0.057 | 0.01 (-0.5,0.53) | 0.26 | 0.26 | 0.046 | 0.01 (-0.5,0.53) | 0.26 | 0.26 | 0.046 |
| A(EM) | -0.01 (-0.33,0.31) | 0.17 | 0.16 | 0.042 | -0.01 (-0.72,0.7) | 0.36 | 0.36 | 0.046 | 3.89 (2.62,5.16) | 0.64 | 0.65 | 1 | 0.00 (-0.23,0.22) | 0.11 | 0.11 | 0.052 | 0.01 (-0.5,0.52) | 0.28 | 0.26 | 0.039 | 0.01 (-0.5,0.51) | 0.26 | 0.26 | 0.05 |
| *MAR dropout* | | | | | | | | | | | | | *MAR dropout* | | | | | | | | | | | |
| B | 0.01 (-0.33,0.34) | 0.17 | 0.17 | 0.039 | 0.01 (-0.82,0.84) | 0.43 | 0.42 | 0.04 | 0.02 (-1.00,1.03) | 0.54 | 0.52 | 0.033 | 0.00 (-0.25,0.25) | 0.13 | 0.13 | 0.044 | 0.02 (-0.59,0.64) | 0.31 | 0.31 | 0.056 | 0.03 (-0.59,0.64) | 0.31 | 0.31 | 0.056 |
| B(EM) | 0.00 (-0.37,0.38) | 0.21 | 0.19 | 0.036 | 0.01 (-0.82,0.84) | 0.43 | 0.42 | 0.047 | 3.89 (2.41,5.38) | 0.82 | 0.76 | 0.999 | 0.00 (-0.27,0.27) | 0.14 | 0.14 | 0.06 | -0.24 (-0.89,0.4) | 0.34 | 0.33 | 0.106 | 0.00 (-0.63,0.63) | 0.31 | 0.32 | 0.055 |
| *Outcome-dependent MNAR dropout* | | | | | | | | | | | | | *Outcome-dependent MNAR dropout* | | | | | | | | | | | |
| C | -0.44 (-0.7,-0.19) | 0.13 | 0.13 | 0.938 | -0.18 (-0.85,0.49) | 0.34 | 0.34 | 0.075 | -0.52 (-1.13,0.08) | 0.31 | 0.31 | 0.377 | -0.32 (-0.53,-0.10) | 0.11 | 0.11 | 0.836 | -0.26 (-0.68,0.16) | 0.22 | 0.21 | 0.232 | -0.26 (-0.68,0.16) | 0.21 | 0.21 | 0.236 |
| C(EM) | -0.99 (-1.26,-0.72) | 0.14 | 0.14 | 1 | -0.80 (-0.85,0.49) | 0.33 | 0.32 | 0.706 | 1.07 (0.24,1.91) | 0.41 | 0.43 | 0.75 | -0.64 (-0.86,-0.42) | 0.11 | 0.11 | 1 | 0.04 (-0.41,0.49) | 0.24 | 0.23 | 0.037 | -0.03 (-0.47,0.42) | 0.23 | 0.23 | 0.047 |
| *Non-outcome-dependent MNAR dropout* | | | | | | | | | | | | | *Non-outcome-dependent MNAR dropout* | | | | | | | | | | | |
| D | -0.27 (-0.6,0.06) | 0.16 | 0.17 | 0.389 | 0.02 (-0.79,0.84) | 0.42 | 0.42 | 0.05 | 0.15 (-0.82,1.12) | 0.5 | 0.5 | 0.059 | -0.28 (-0.52,-0.04) | 0.12 | 0.12 | 0.632 | 0.15 (-0.39,0.68) | 0.28 | 0.27 | 0.067 | 0.15 (-0.39,0.69) | 0.28 | 0.27 | 0.069 |
| D(EM) | -0.28 (-0.63,0.08) | 0.2 | 0.18 | 0.262 | 0.00 (-0.82,0.82) | 0.42 | 0.42 | 0.045 | 3.99 (2.57,5.41) | 0.77 | 0.73 | 1 | -0.28 (-0.53,-0.03) | 0.13 | 0.13 | 0.575 | -0.04 (-0.61,0.52) | 0.31 | 0.29 | 0.042 | 0.12 (-0.43,0.68) | 0.28 | 0.28 | 0.074 |

M2 *MNAR dropout dependent on $B$, $X$, and $Y$*

- $\mathsf{P}(R = 1|Y, X, A, B) = \exp(2.8 + 2.1X + 2.1B + 2.1Y)/(1 + \exp(2.8 + 2.1X + 2.1B + 2.1Y))$

**Simulation size:** For each scenario, 1000 datasets were simulated. Simulation quality was verified by checking of mean SEs of the CCA and variance difference estimates were comparable to the Monte Carlo SEs (see 'Performance measures')

**Estimands**
*Primary:* Reported in main text, Table 2, and in appendix, Table C.1.

- Bias of the complete case analysis (CCA) treatment effect estimate, regressing outcome, $Y$, on treatment, $X$, and covariate $A$, with 95% CI
- Outcome variance difference adjusted for $A$ , with 95% CI
- Bias of the MI treatment effect estimate, regressing outcome, $Y$, on treatment, $X$, and covariate $A$, in datasets with outcomes imputed with an imputation model including $Y$, $X$, $A$ and $B$, with 95% CI.
- Outcome variance difference adjusted for $A$, estimated in datasets with outcomes imputed with an imputation model including $Y$, $X$, $A$ and $B$, with 95% CI.

*Secondary:* Reported in appendix, Table C.1.

- 95% coverage* of all four measures listed above
- Standard error (SE) of all four measures listed above
- Monte Carlo SE (MCSE) of all four measures listed above

(* The 95% coverage refers to the proportion of times the estimate was excluded from the 95% CI. If the true estimate is 0, this should come to 0.05)

**Methods**

- CCA estimator
- Multiple imputation, performed using $R$ software package 'mice'
- Studentized Breusch-Pagan test (for estimating the variance difference across groups after dropout in the observed data, and in the imputed datasets)
- 95% CIs were calculated using the Monte Carlo Standard error (MCSE)

**Performance measures**: Simulation quality was checked by seeing if the estimate SEs were comparable to the MCSEs

#### C.2 Simulation results

**Table C.1:** Bias of complete case analysis (CCA) and multiple imputation (MI) treatment effect estimates and variance differences (VD) with 95% confidence intervals (CIs), for data ($N = 1000$, $N = 10000$) simulated according to DAGS M1 and M2 (Figure 5 in the main text). M1) $Y$ is a function of $A$, $B$ and treatment, $X$, with dropout dependent on $A$ and $B$; M2) Analogous to M1, with dropout additionally dependent on $Y$. Shown is the CCA treatment effect estimate, conditional on $A$, with corresponding VD, alongside the MI treatment effect estimate and VD ($\text{VD}_{MI}$), estimated conditional on $A$, with both $A$ and $B$ included in the imputation model, with for each the 95% CI, standard error (SE), Monte Carlo standard error (MCSE) and proportion of 95% CIs excluding the null ($p_{95\%\text{CI}}$. Dropout proportions in the comparator and intervention group are denoted $p_0$ and $p_1$, respectively.

| | *Observed data* | | *Imputed data* | | *Dropout* | |
|---|---|---|---|---|---|---|
| | Bias (95% CI) | $\text{VD}_f$ (95% CI) | Bias (95% CI) | $\text{VD}_{f(MI)}$ (95% CI) | $p_0$ | $p_1$ |
| M1 | -0.35 (-0.68,-0.02) | 0.25 (-0.82,1.33) | -0.02 (-0.37,0.33) | 0.08 (-0.82,0.97) | 0.18 | 0.37 |
| M2 | -0.56 (-0.85,-0.28) | 0.75 (-0.04,1.54) | -0.49 (-0.75,-0.23) | 0.77 (0.14,1.4) | 0.19 | 0.36 |
